# Sex Differences in Social, Behavioral, and Metabolic Risk Factors for Cardiovascular Disease Mortality among US Adults

**DOI:** 10.1101/2025.11.20.25340714

**Authors:** Ling Tian, Siyi Geng, Xingyan Li, Byron C. Jaeger, Kirsten S. Dorans, Jing Chen, Amanda H. Anderson, Paul K. Whelton, Marie Krousel-Wood, Jiang He, Joshua D. Bundy

## Abstract

**Background:** Prevalence and associations of risk factors for cardiovascular disease (CVD) may differ between women and men, but previous findings are inconclusive. We investigated the simultaneous contributions of social determinants of health (SDOH), behavioral factors, and metabolic factors to CVD mortality among women and men in the US general population.

**Methods:** We included 50,808 participants aged ≥20 years from the National Health and Nutrition Examination Survey 1999-2018, linked to the National Death Index for CVD mortality follow-up through December 31, 2019. SDOH, behavioral factors, and metabolic factors were collected in each survey cycle. We estimated the simultaneous impact of SDOH, behavioral factors, and metabolic factors on CVD mortality in women and men using average population attributable fractions (PAFs), which consider both the prevalence and strength of association of all risk factors with CVD mortality in one model.

**Results:** During a mean 9.4-year follow-up, 2,589 CVD deaths were recorded (1,140 in women; 1,449 in men). Women had higher prevalence of unemployment and central obesity than men. Men had higher prevalence of no regular healthcare access, current smoking, heavy alcohol drinking, unhealthy diet, and high cholesterol than women. In fully adjusted models, associations of risk factors with CVD mortality were mostly similar among women and men. However, unemployment was more strongly associated with CVD mortality among men (hazard ratio [95% CI], 1.97 [1.62-2.39]) compared with women (1.19 [1.00-1.43; p-value for interaction <0.001). In those older than age 60, sleep duration <6 or >8 hours/day was independently associated with CVD mortality among women but not men. Hypertension (14.3%), albuminuria (10.0%), low family income-to-poverty ratio (8.7%), leisure-time physical inactivity (6.6%), and diabetes (6.3%) were top population-level contributors to CVD mortality in men, while leisure-time physical inactivity (14.7%), albuminuria (11.7%), low family income-poverty ratio (10.0%), low estimated glomerular filtration rate (8.9%), and diabetes (6.5%) were top contributors among women.

**Conclusions:** Although the prevalence of CVD risk factors varied by sex, magnitudes of association with CVD mortality were mostly similar among women and men. SDOH and behavioral risk factors collectively accounted for a third of CVD deaths in both men and women living in the US.

**Clinical Perspective:** *What Is New?:*

- Associations of a wide array of social determinants of health, behavioral factors, and metabolic factors with CVD mortality are similar among men and women in the US. However, prevalence of these risk factors and associated population attributable fractions vary by sex.
- Hypertension, lower kidney function, and leisure-time physical inactivity are top population-level contributors to CVD mortality among men, while leisure-time physical inactivity, low kidney function and diabetes are top population-level contributors among women.

*What Are the Clinical Implications?:*

- Low kidney function showed the largest absolute mortality rate in both men and women compared to other risk factors, and increased efforts targeting prevention of kidney disease are warranted.
- Sex-specific strategies may be warranted to target the primordial prevention of social determinants of health and behavioral factors to reduce the burden of CVD mortality.

## Introduction

Cardiovascular disease (CVD) remains the leading cause of death among both women and men in the US, and declines in CVD mortality are plateauing.^1,2^ In general, men are at higher risk for CVD than women across all age groups, although these differences in sex-specific rates decrease with age.^3,4^ Most previous reports have focused on sex differences in metabolic and certain lifestyle risk factors.^3,5–10^ A nationally representative study of US adults found systolic blood pressure (BP) and total cholesterol were lower among women than men before age 50, but higher after age 50.^11^ Previous studies identified that elevated BP, diabetes and smoking were associated with CVD events in both sexes, but to a larger extent in women.^3–6,12^ However, sex differences in the effects of diet, stress and alcohol drinking on CVD events are inconclusive.^3,4,6^

Although some biological factors underlying CVD risk are distinct between men and women, there is a complex interplay among modifiable social, behavioral, lifestyle, and metabolic factors.^13^ For example, a growing body of evidence supports the critical role that psychological health and social determinants of health (SDOH) play in CVD.^14^ However, the extent to which combined SDOH, behavioral factors, and metabolic factors differ between women and men, and how these factors affect population-level risk for CVD mortality, have not been determined in the US general population.^15^ Investigating the independent, direct associations of SDOH, behavioral factors, and metabolic factors on CVD mortality by sex could aid in the development of optimal strategies for reducing the burden of CVD among all adults.

The aims of the current study were to (1) quantify sex-specific levels of a comprehensive array of SDOH, behavioral factors, and metabolic factors; (2) assess the independent associations of these risk factors with CVD mortality among men and women; and (3) estimate the extent to which these differences in risk factors can explain US population-level sex differences in CVD mortality. To evaluate these aims, we used data from a representative sample of US adults studied in the National Health and Nutrition Examination Surveys (NHANES) 1999 to 2018 with linkage to the National Death Index (NDI), with follow-up for CVD mortality through 2019.

## Methods

### Study Design and Participants

NHANES comprises a series of cross-sectional, nationally representative samples of the civilian, noninstitutionalized US population using stratified, multistage probability sampling methods in 2-year cycles since 1999.^16^ We included 10 cycles conducted between 1999 to 2000 and 2017 to 2018. The present study included 50,808 non-pregnant participants aged ≥20 years with unique identifiers sufficient for linkage to the NDI. We excluded 122 individuals due to insufficient identifiers. All participants provided written informed consent, and the Ethics Review Board of the National Center for Health Statistics approved all protocols.

### Data Collection and Risk Factor Assessment

Each participant completed a household interview, responded to additional questionnaires, and underwent physical examination and laboratory testing in a mobile examination center (MEC) using standardized protocols, which were used throughout the whole data collection process.^17^ We included 22 risk factors across three perspectives which have previously been associated with CVD: SDOH, behavioral factors, and metabolic factors.^2,4,18^ Detailed definitions are available in **Supplemental Table 1**. SDOH and behavioral risk factors were ascertained from questionnaire interview while metabolic factors were ascertained using physical and laboratory testing.

We included nine SDOH: employment status, family income to poverty ratio, food security, high school education, regular healthcare access, health insurance status, home ownership, crowded housing, and marital status.^19–21^ Employment was defined as currently working or if not working, being a student or retired. Family income to poverty ratio was calculated by dividing family income by a poverty threshold related to family size and adjusted for geographic differences and inflation. Food security was assessed using the validated US Food Security Module.^22^

We included six behavioral risk factors, including psychological and lifestyle factors: current smoking, heavy alcohol drinking, low diet quality, leisure-time physical inactivity, sleep duration <6 or >8 hours per day, and depression. Heavy alcohol drinking was defined as more than 14 drinks per week for men, and 7 for women.^23^ Responses to 24-hour dietary recalls were used to calculate the Healthy Eating Index-2015 (HEI-2015), ranging from 0 to 100 with higher scores indicating greater consistency of the diet with the Dietary Guidelines for Americans 2015-2020.^24–26^ Because questionnaires assessing physical activity changed after 2004, only leisure-time physical activity was assessed in the current study.^18^ Depression was measured using the Patient Health Questionnaire-9 (PHQ-9), with scores ≥10 indicating depression.^27^

We included seven metabolic risk factors: obesity, central obesity, hypertension, diabetes, high cholesterol, urinary albumin-to-creatinine ratio and estimated glomerular filtration rate (eGFR). Height, weight, and waist circumference were measured during the physical examination.

Systolic BP was calculated as the mean of 3 readings obtained using standardized procedures.^17^ HDL cholesterol, total cholesterol, triglycerides, plasma glucose, and glycated hemoglobin (HbA1c) were measured using blood collected at the MEC using standardized protocols.^17^ Body mass index (BMI) was calculated as the weight (measured in kilograms) divided by height (measured in meters) squared. Obesity was defined as BMI ≥30 kg/m^2^, and central obesity was defined as waist circumference ≥102 cm for men and ≥88 cm for women.^28^ Antihypertensive, lipid lowering, and antidiabetic medication use were self-reported. Hypertension was defined as systolic BP ≥130 mmHg and/or diastolic BP ≥80 mmHg or use of antihypertensive medications.^29^ Diabetes was defined as fasting glucose ≥126mg/dL or HbA1c ≥6.5%, or diagnosed diabetes.^30^ High cholesterol was defined as ratio of total to HDL cholesterol ≥5.^31^ Urinary albumin and creatine were collected with random spot urine samples. eGFR was calculated using the 2021 Chronic Kidney Disease Epidemiology Collaboration (CKD-EPI) creatine-based equation.^32^

### Outcome Ascertainment

The mortality status of each NHANES participant was matched and linked to the NDI through December 31, 2019. International Classification of Diseases, Tenth Revision (ICD-10) codes were used to identify participants for whom heart disease (codes I00-I09, I11, I13, I20-I51) or stroke (codes I60-I69) were listed as the underlying cause of death. Participants continued their follow-up until death attributable to CVD, with censoring at the time of death from non-CVD causes or end of follow-up. Participants not matched with a death record were considered alive throughout the follow-up period. A detailed description of the matching methods is available elsewhere.^33^

### Statistical Analysis

To obtain nationally representative estimates, all analyses in this study account for the complex, multistage, probability design of NHANES using the *survey* package in R.^34^ To estimate average populational attributable fractions (PAFs), risk factors were dichotomized based on common clinical cut-off points, or the median if a linear association with CVD mortality was identified. Since comprehensive information is collected in NHANES, it is reasonable to assume that data are missing at random.^35^ Therefore, multiple imputation by chained equations was used to address partially missing and systematically missing data for specific cycles (sleep and depression data were missing in NHANES 1999-2004) in the primary analysis.^36,37^ Ten imputed datasets were generated and results from each dataset were combined into final estimates for statistical inference using Rubin’s Rules.^38^

Baseline characteristics of participants (proportion and 95% confidence interval [CI] for categorical variables; mean and 95% CI for continuous variables) were summarized in the overall population and compared between men and women. Interactions between each risk factor and age group were tested, which identified several statistically significant interactions.

Therefore, results were also presented stratified by age (<60, and ≥60 years), which also sought to capture risk profiles among women before and after the menopause transition.^5,6,39^ Incidence rates were calculated for each risk factor by sex and age subgroup and in the overall population as the number of CVD deaths per 1,000 person-years. Cox proportional hazards regression using age as the time scale with adjustment for left truncation was used to investigate the independent associations of SDOH, behavioral factors, and metabolic factors with CVD mortality.^40^ The models were stratified by birth cohort to adjust for potential secular trends. The primary model included all risk factors and their interactions with sex, adjusted for race/ethnicity. The assumption of proportionality of hazards was assessed by visual inspection of Schoenfeld residuals and no violations were detected.

Average PAFs were estimated to quantify the extent to which US population-level CVD mortality is explained by each risk factor, separately in women and men. PAFs and 95% CIs were estimated for SDOH, behavioral factors, and metabolic factors significantly associated with CVD mortality in the fully adjusted Cox proportional models by each sex and age subgroup.

Risk factors not statistically significantly associated with CVD mortality were included as covariates for adjustment in PAF estimation. Average PAFs were estimated using methods developed by Edie and Gefeller and were adapted from the R package *averisk.*^41,42^

All analyses were conducted using SAS Statistical Software, version 9.4 (SAS Institute Inc) and R software, version 4.2.2 (R Foundation). All statistical tests were 2 sided, and p<0.05 was considered statistically significant. Given the multiplicity of comparisons, results should be interpreted as exploratory.

## Results

Among 50,808 NHANES participants aged ≥20 years, the mean survey-weighted age was 47.3 years and 50.5% were women (**Table 1**). Family income to poverty ratio <300%, unhealthy diet, leisure-time physical inactivity, central obesity and hypertension were highly prevalent in both men and women. Compared with men, women were more likely to be unemployed, have family income to poverty ratio <300%, not be married nor living with a partner, have depression, leisure-time physical inactivity, sleep <6 or >8 hours per day, have obesity and central obesity, and have albuminuria and eGFR <60 ml/min/1.73 m^2^. Men were more likely to have no regular health care access, current smoking, heavy alcohol drinking, unhealthy diet, and high cholesterol compared with women. Similar patterns were observed after stratifying by age (**Figure 1 and Supplemental Table 2**). Furthermore, the prevalence of not being married nor living with a partner, leisure-time physical inactivity, central obesity, hypertension, diabetes and eGFR <60 ml/min/1.73 m^2^ increased substantially for both men and women, and particularly for women aged ≥60 years.

**Figure 1.**
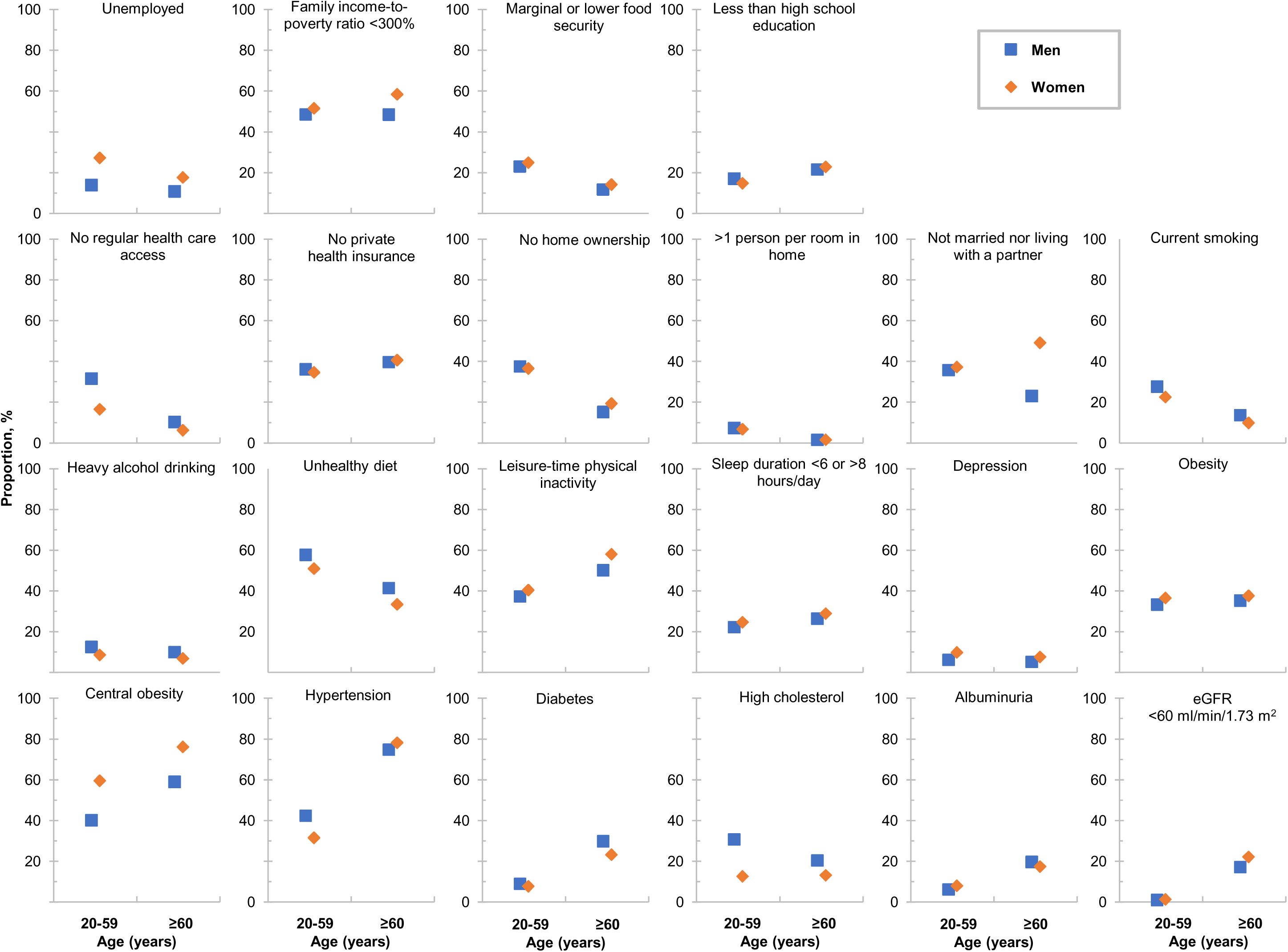
Prevalence of Social Determinants of Health, Behavioral Factors, and Metabolic Factors by Sex and Age Group * All data points in the figure are weighted proportions accounting for multiple imputation, and 95% CIs are shown in Supplemental Table 2 because they were too narrow to be seen clearly on the figure. Employment is defined as currently working, student, or retired. Food insecurity is assessed with the 10-item adult US Food Security Survey Module with zero affirmative responses indicating high food security and ≥1 affirmative responses indicating marginal or lower food security. Heavy alcohol drinking is defined as consuming more than 14 drinks per week for men, and 7 for women. The Healthy Eating Index (HEI) uses a scoring system to assess how well a set of foods aligns with key recommendations of the Dietary Guidelines for Americans 2015-2020. The scores range from 0 to 100 with a higher score indicating greater consistency of the diet with the Dietary Guidelines for Americans. Median HEI score was 52 among NHANES 1999-2018 participants. Depression was assessed with the Patient Health Questionnaire-9 (PHQ-9) with scores ≥10 indicating depression. Obesity is defined as body mass index ≥30 kg/m2; central obesity is defined as waist circumference ≥102 cm for men and ≥88 cm for women; hypertension is defined as systolic blood pressure ≥130 mmHg and/or diastolic blood pressure ≥80 mmHg or use of antihypertensive medications; and diabetes is defined as fasting glucose ≥126mg/dL or hemoglobin A1c ≥6.5% or diagnosed diabetes.

**Table 1.** Baseline Characteristics and Risk Factors Among US Men and Women, NHANES 1999-2018.

| Characteristics <sup>*</sup> | All US adults<br>(N = 50,808) | Men<br>(N = 25,143) | Women<br>(N = 25,665) | P-value |
| --- | --- | --- | --- | --- |
| Age in years, mean (95% CI) | 47.3 (47.0, 47.7) | 46.3 (45.9, 46.6) | 48.3 (47.9, 48.7) | <0.001 |
| Race/ethnicity, % (95% CI) |  |  |  | <0.001 |
| Non-Hispanic Black | 11.2 (10.1, 12.3) | 10.4 (9.4, 11.5) | 11.9 (10.7, 13.1) |  |
| Hispanic | 13.7 (12.2, 15.2) | 14.3 (12.8, 15.9) | 13.1 (11.6, 14.6) |  |
| Non-Hispanic White | 68.2 (66.2, 70.2) | 68.3 (66.3, 70.3) | 68.1 (66.0, 70.2) |  |
| Others | 6.9 (6.3, 7.5) | 6.9 (6.2, 7.5) | 6.9 (6.3, 7.5) |  |
| Social determinants of health, % (95% CI) <sup>†</sup> |  |  |  |  |
| Unemployed | 19.1 (18.4, 19.8) | 13.3 (12.5, 14.0) | 24.6 (23.7, 25.5) | <0.001 |
| Family income-to-poverty ratio <300% | 51.1 (49.7, 52) | 48.6 (47.1, 50.1) | 53.5 (52.0, 55.0) | <0.001 |
| Marginal or lower food security | 21.2 (20.4, 22.1) | 20.4 (19.5, 21.4) | 22.0 (21.1, 23.0) | <0.001 |
| Less than high school education | 17.6 (16.7, 18.5) | 18.2 (17.2, 19.2) | 17.0 (16.2, 17.9) | 0.001 |
| No regular health care access | 20.1 (19.3, 20.8) | 26.7 (25.7, 27.6) | 13.8 (13.1, 14.5) | <0.001 |
| No private health insurance | 36.7 (35.6, 37.8) | 37.0 (35.8, 38.2) | 36.4 (35.2, 37.6) | 0.20 |
| No home ownership | 32.1 (30.8, 33.5) | 32.4 (31.0, 33.8) | 31.9 (30.5, 33.2) | 0.21 |
| >1 person per room in home | 5.7 (5.1, 6.3) | 6.1 (5.4, 6.7) | 5.3 (4.8, 5.9) | <0.001 |
| Not married nor living with a partner | 36.8 (35.9, 37.7) | 32.8 (31.8, 33.9) | 40.6 (39.6, 41.6) | <0.001 |
| Behavioral factors, % (95% CI) <sup>‡</sup> |  |  |  |  |
| Current smoking | 21.7 (21.0, 22.4) | 24.4 (23.5, 25.3) | 19.1 (18.3, 19.9) | <0.001 |
| Heavy alcohol drinking | 10.1 (9.6, 10.6) | 12.0 (11.5, 12.6) | 8.2 (7.6, 8.9) | <0.001 |
| Unhealthy diet | 50.0 (48.9, 51.1) | 54.0 (52.9, 55.2) | 46.2 (44.8, 47.5) | <0.001 |
| Leisure-time physical inactivity | 42.9 (41.8, 44.0) | 40.4 (39.1, 41.6) | 45.3 (44.1, 46.6) | <0.001 |
| Sleep duration <6 or >8 hours/day | 24.6 (24.0, 25.2) | 23.3 (22.5, 24.1) | 25.9 (25.1, 26.6) | <0.001 |
| Depression | 7.7 (7.3, 8.1) | 6.0 (5.6, 6.5) | 9.3 (8.8, 9.8) | <0.001 |
| Metabolic factors, % (95% CI) <sup>§</sup> |  |  |  |  |
| Obesity | 35.4 (34.6, 36.2) | 33.9 (32.8, 34.9) | 36.9 (36.0, 37.8) | <0.001 |
| Central obesity | 54.6 (53.6, 55.6) | 44.6 (43.4, 45.7) | 64.2 (63.1, 65.2) | <0.001 |
| Hypertension | 47.1 (46.3, 47.9) | 49.9 (48.9, 50.9) | 44.4 (43.4, 45.3) | <0.001 |
| Diabetes | 12.9 (12.5, 13.4) | 13.8 (13.2, 14.4) | 12.1 (11.5, 12.7) | <0.001 |
| High cholesterol | 20.4 (19.8, 21.0) | 28.4 (27.5, 29.3) | 12.8 (12.2, 13.4) | <0.001 |
| Albuminuria | 10.0 (9.6, 10.4) | 9.3 (8.9, 9.8) | 10.7 (10.2, 11.2) | <0.001 |
| eGFR <60 ml/min/1.73 m<sup>2</sup> | 6.0 (5.7, 6.3) | 4.9 (4.5, 5.2) | 7.1 (6.7, 7.5) | <0.001 |

Abbreviations: CI, confidence interval; eGFR, estimated glomerular filtration rate; NHANES, National Health and Nutrition Examination Survey.
\* Sample sizes are unobserved frequencies while the other numbers in the table are weighted means (95% CI) or percentages (95% CI) after multiple imputation.
† Employment is defined as currently working, student, or retired. Food insecurity is assessed with the 10-item adult US Food Security Survey Module with zero affirmative responses indicating high food security and ≥1 affirmative responses indicating marginal or lower food security.
‡ Heavy alcohol drinking is defined as consuming more than 14 drinks per week for men, and 7 for women. The Healthy Eating Index (HEI) uses a scoring system to assess how well a set of foods aligns with key recommendations of the Dietary Guidelines for Americans 2015-2020. The scores range from 0 to 100 with a higher score indicating greater consistency of the diet with the Dietary Guidelines for Americans. Median HEI score was 52 among NHANES 1999-2018 participants; Depression was assessed with the Patient Health Questionnaire-9 (PHQ-9) with scores ≥10 indicating depression.
§ Obesity is defined as body mass index ≥30 kg/m<sup>2</sup>; central obesity is defined as waist circumference ≥102 cm for men and ≥88 cm for women; hypertension is defined as systolic blood pressure ≥130 mmHg and/or diastolic blood pressure ≥80 mmHg or use of antihypertensive medications; and diabetes is defined as fasting glucose ≥126mg/dL or hemoglobin A1c ≥6.5% or diagnosed diabetes.

A total of 2,589 CVD deaths were recorded (men, 1,449; women, 1,140) during a mean 9.4 years of follow-up. The age-adjusted CVD mortality rates (95% CIs) per 100,000 person-years were 475 (429-527) and 358 (320-402) among men and women, respectively. Absolute CVD mortality rates within most risk factor groups were similar among men and women, overall and by age subgroups (**Table 2**). However, among those who were unemployed, the CVD mortality rate was higher among men (6.0 deaths per 1,000 person-years) compared with women (2.9 deaths per 1,000 person-years). Conversely, among those not married nor living with a partner, CVD mortality rates were lower among men (4.0 deaths per 1,000 person-years) compared with women (5.4 deaths per 1,000 person-years). Additionally, CVD mortality rates were higher among men with albuminuria or eGFR <60 ml/min/1.73 m^2^, but lower with high cholesterol compared with women. In those 60 years of age and older, absolute CVD mortality rates were consistently higher among men compared with women for all risk factors except crowded housing and high cholesterol.

**Table 2.** Absolute Risks of Cardiovascular Disease Mortality Among US Men and Women by Age Group, NHANES 1999-2018 *.

| Risk Factors | Mortality Rate, events per 1,000 person-years |  |  |  |  |  |
| --- | --- | --- | --- | --- | --- | --- |
|  | All US adults |  | Age <60 years |  | Age ≥60 years |  |
|  | Men | Women | Men | Women | Men | Women |
| **Social determinants of health <sup>†</sup>** |  |  |  |  |  |  |
| Employment status |  |  |  |  |  |  |
| Employed, student, or retired | 3.5 | 3.5 | 0.8 | 0.5 | 15.0 | 12.6 |
| Unemployed | 6.0 | 2.9 | 3.8 | 1.3 | 19.9 | 11.6 |
| Family income-to-poverty ratio |  |  |  |  |  |  |
| ≥300% | 2.7 | 1.7 | 0.9 | 0.4 | 10.5 | 7.0 |
| <300% | 5.0 | 4.8 | 1.6 | 1.0 | 21.2 | 16.2 |
| Food security |  |  |  |  |  |  |
| Full security | 3.8 | 3.5 | 1.0 | 0.6 | 15.1 | 12.3 |
| Marginal, low, or very low security | 3.5 | 2.9 | 1.9 | 1.2 | 18.7 | 13.3 |
| Education level |  |  |  |  |  |  |
| High school graduate or higher | 3.2 | 2.6 | 1.0 | 0.5 | 13.4 | 10.4 |
| Less than high school | 6.4 | 6.8 | 2.0 | 1.6 | 22.7 | 18.5 |
| Regular health care access |  |  |  |  |  |  |
| At least one regular health care facility | 4.5 | 3.6 | 1.3 | 0.7 | 15.6 | 12.4 |
| None or emergency room | 1.7 | 1.8 | 0.9 | 0.6 | 13.5 | 12.2 |
| Health insurance status |  |  |  |  |  |  |
| Private | 3.1 | 2.6 | 0.9 | 0.5 | 13.6 | 10.7 |
| Government or none | 5.0 | 4.8 | 1.7 | 1.2 | 18.5 | 15.0 |
| Home ownership |  |  |  |  |  |  |
| Own home | 4.2 | 3.5 | 1.1 | 0.6 | 14.4 | 11.4 |
| Rent home or other arrangement | 2.9 | 3.0 | 1.4 | 0.9 | 21.8 | 17.1 |
| Number of persons per room in home |  |  |  |  |  |  |
| ≤1 person | 3.9 | 3.5 | 1.2 | 0.7 | 15.5 | 12.4 |
| >1 person | 1.4 | 1.3 | 1.0 | 0.7 | 8.8 | 9.4 |
| Marital status |  |  |  |  |  |  |
|  | All US adults |  | Age <60 years |  | Age ≥60 years |  |
|  | Men | Women | Men | Women | Men | Women |
| Married or living with a partner | 3.7 | 2.0 | 1.0 | 0.5 | 13.4 | 8.0 |
| Not married nor living with a partner | 4.0 | 5.4 | 1.6 | 1.0 | 23.7 | 17.6 |
| **Behavioral factors <sup>‡</sup>** |  |  |  |  |  |  |
| Smoking |  |  |  |  |  |  |
| Former or never smoking | 4.0 | 3.6 | 0.9 | 0.5 | 15.1 | 12.4 |
| Current smoking | 3.3 | 2.4 | 1.9 | 1.3 | 17.4 | 12.0 |
| Alcohol drinking |  |  |  |  |  |  |
| Moderate or no alcohol drinking | 3.9 | 3.5 | 1.1 | 0.7 | 15.9 | 12.8 |
| Heavy alcohol drinking | 3.2 | 1.9 | 1.6 | 0.6 | 11.7 | 7.1 |
| Diet quality |  |  |  |  |  |  |
| HEI ≥52 | 4.3 | 3.8 | 1.0 | 0.5 | 14.6 | 11.7 |
| HEI <52 | 3.4 | 2.8 | 1.4 | 0.9 | 16.7 | 13.7 |
| Leisure-time physical activity |  |  |  |  |  |  |
| Any | 2.6 | 1.7 | 0.8 | 0.5 | 11.8 | 7.4 |
| No | 5.9 | 5.6 | 1.9 | 1.1 | 19.8 | 16.6 |
| Sleep duration |  |  |  |  |  |  |
| 6-8 hours per day | 3.5 | 3.0 | 1.1 | 0.6 | 14.8 | 11.3 |
| <6 or >8 hours per day | 4.8 | 4.4 | 1.8 | 0.9 | 17.4 | 15.6 |
| Depression |  |  |  |  |  |  |
| PHQ-9 <10 | 3.7 | 3.3 | 1.1 | 0.6 | 15.2 | 12.1 |
| PHQ-9 ≥10 | 4.8 | 4.3 | 2.3 | 1.7 | 19.9 | 15.9 |
| **Metabolic factors <sup>§</sup>** |  |  |  |  |  |  |
| Obesity |  |  |  |  |  |  |
| No | 3.4 | 3.2 | 1.0 | 0.5 | 15.3 | 12.7 |
| Yes | 4.5 | 3.7 | 1.8 | 1.1 | 15.7 | 11.8 |
| Central obesity |  |  |  |  |  |  |
|  | All US adults |  | Age <60 years |  | Age ≥60 years |  |
|  | Men | Women | Men | Women | Men | Women |
| No | 2.6 | 2.1 | 0.8 | 0.4 | 14.9 | 12.5 |
| Yes | 5.4 | 4.1 | 1.8 | 0.9 | 15.8 | 12.3 |
| Hypertension |  |  |  |  |  |  |
| No | 1.4 | 0.8 | 0.5 | 0.3 | 11.0 | 7.1 |
| Yes | 6.3 | 6.8 | 2.2 | 1.7 | 16.9 | 13.9 |
| Diabetes |  |  |  |  |  |  |
| No | 2.8 | 2.5 | 0.9 | 0.4 | 13.5 | 10.9 |
| Yes | 11.7 | 10.6 | 4.7 | 4.1 | 20.8 | 17.9 |
| High cholesterol |  |  |  |  |  |  |
| No | 3.9 | 3.1 | 1.0 | 0.6 | 15.6 | 11.9 |
| Yes | 3.4 | 4.8 | 1.6 | 1.6 | 15.0 | 15.2 |
| Albumin-to-creatinine ratio |  |  |  |  |  |  |
| <30 mg/g | 2.8 | 2.4 | 0.9 | 0.5 | 12.4 | 9.5 |
| ≥30 mg/g | 16.1 | 12.3 | 6.4 | 2.8 | 31.3 | 30.0 |
| Estimated glomerular filtration rate |  |  |  |  |  |  |
| ≥60 ml/min/1.73 m<sup>2</sup> | 2.9 | 2.2 | 1.1 | 0.6 | 12.3 | 8.9 |
| <60 ml/min/1.73 m<sup>2</sup> | 29.1 | 24.0 | 9.4 | 9.5 | 35.1 | 27.1 |

Abbreviations: NHANES, National Health and Nutrition Examination Survey.
\* All numbers in the table are weighted to the US population and account for multiple imputation.
† Employment is defined as currently working, student, or retired. Food insecurity is assessed with the 10-item adult US Food Security Survey Module with zero affirmative responses indicating high food security and ≥1 affirmative responses indicating marginal or lower food security.
‡ Heavy alcohol drinking is defined as consuming more than 14 drinks per week for men, and 7 for women. The Healthy Eating Index (HEI) uses a scoring system to assess how well a set of foods aligns with key recommendations of the Dietary Guidelines for Americans 2015-2020. The scores range from 0 to 100 with a higher score indicating greater consistency of the diet with the Dietary Guidelines for Americans. Median HEI score was 52 among NHANES 1999-2018 participants; Depression was assessed with the Patient Health Questionnaire-9 (PHQ-9) with scores ≥10 indicating depression.
§ Obesity is defined as body mass index ≥30 kg/m<sup>2</sup>; central obesity is defined as waist circumference ≥102 cm for men and ≥88 cm for women; hypertension is defined as systolic blood pressure ≥130 mmHg and/or diastolic blood pressure ≥80 mmHg or use of antihypertensive medications; and diabetes is defined as fasting glucose ≥126mg/dL or hemoglobin A1c ≥6.5% or diagnosed diabetes.

In minimally adjusted models stratified by birth cohort and adjusted for race and ethnicity, most risk factors were significantly associated with higher CVD mortality in both men and women (**Supplemental Table 3**). Magnitudes of association were mostly similar in men and women.

However, unemployment, marginal or lower food security, and not owning a home were associated with higher risk of CVD mortality among men than women; leisure-time physical inactivity and high cholesterol were associated with higher risk among women than men. After including all risk factors in one fully adjusted model, unemployment was significantly associated with comparatively higher risk of CVD mortality among men than women (HR [95% CI], 1.97 [1.62-2.39] among men, 1.19 [1.00-1.43] among women, P_interaction_, <0.001); leisure-time physical inactivity was significantly associated with lower risk of CVD mortality among men than women: (1.20 [1.03-1.40] among men, 1.44 [1.21-1.72] among women, P_interaction_, 0.16) **(Figure 2 and Supplemental Table 3)**. However, the association between marginal or lower food security with CVD mortality became non-significant in both men and women, not owning a home became non-significant in women, and high cholesterol became non-significant in men.

**Figure 2.**
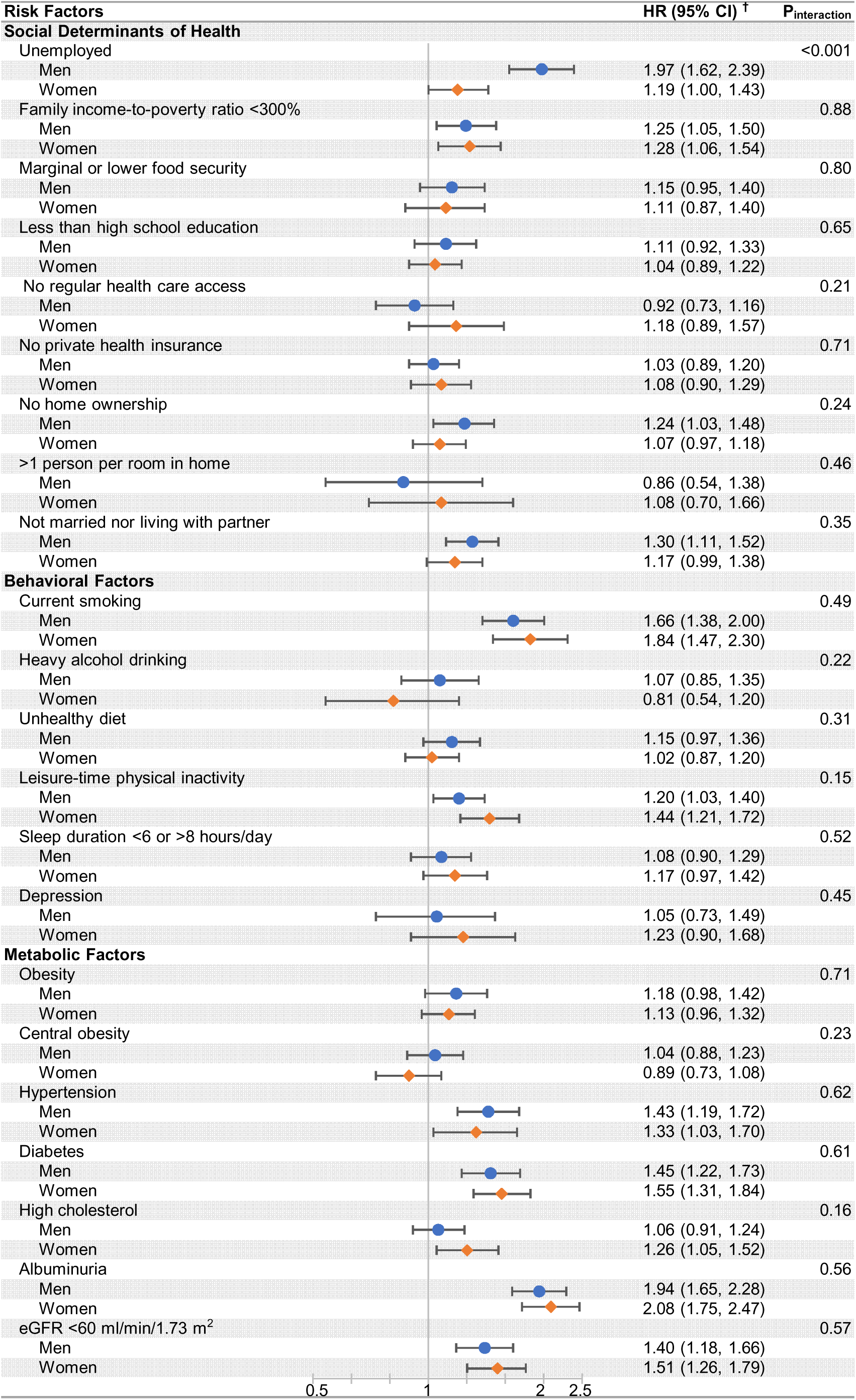
Multivariable-adjusted Associations of Risk Factors with Cardiovascular Disease Mortality by Sex Among All US Adults All numbers in the table are weighted to the US population and account for multiple imputation. Abbreviations: CI, confidence interval; HEI, healthy eating index; eGFR, estimated glomerular filtration rate. ^†^ Age is treated as the time scale and models are stratified by birth cohort and adjusted for sex, race, and all other SDOH, behavioral factors, and metabolic factors listed in the table.

The association between unemployment, and not being married nor living with a partner, with CVD mortality were stronger among men than women in all models across all age groups (**Figures 3**–**4 and Supplemental Tables 4-5**). Depression was associated with higher risk of CVD mortality among women than men aged ≥60 years in minimally adjusted models but became non-significant in fully adjusted models. Current smoking, diabetes and eGFR <60 ml/min/1.73 m^2^ were associated with higher risk of CVD mortality among women than men before age 60, but with similar risk after age 60. Leisure-time physical inactivity, sleep duration <6 or >8 hours per day were associated with similar risk of CVD mortality among women and men before age 60, but higher risk among women than men after age 60. Albuminuria was associated with higher risk of CVD mortality among men than women before age 60, but lower after age 60.

**Figure 3.**
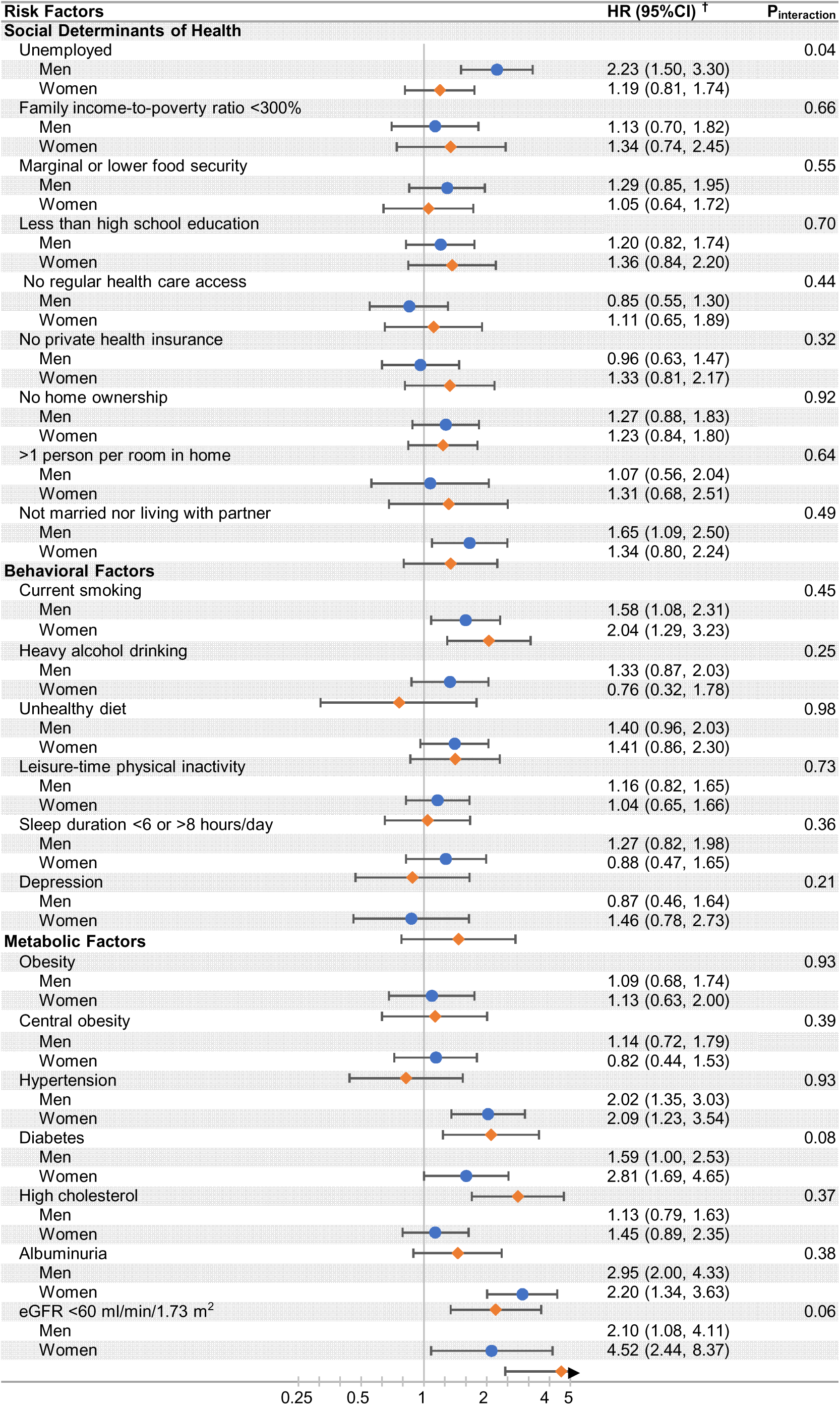
Multivariable-adjusted Associations of Risk Factors with Cardiovascular Disease Mortality by Sex Among Adults Less than 60 Years of Age *All numbers in the table are weighted to the US population and account for multiple imputation. Abbreviations: CI, confidence interval; HEI, healthy eating index; eGFR, estimated glomerular filtration rate. ^†^ Age is treated as the time scale and models are stratified by birth cohort and adjusted for sex, race, and all other SDOH, behavioral factors, and metabolic factors listed in the table.

**Figure 4.**
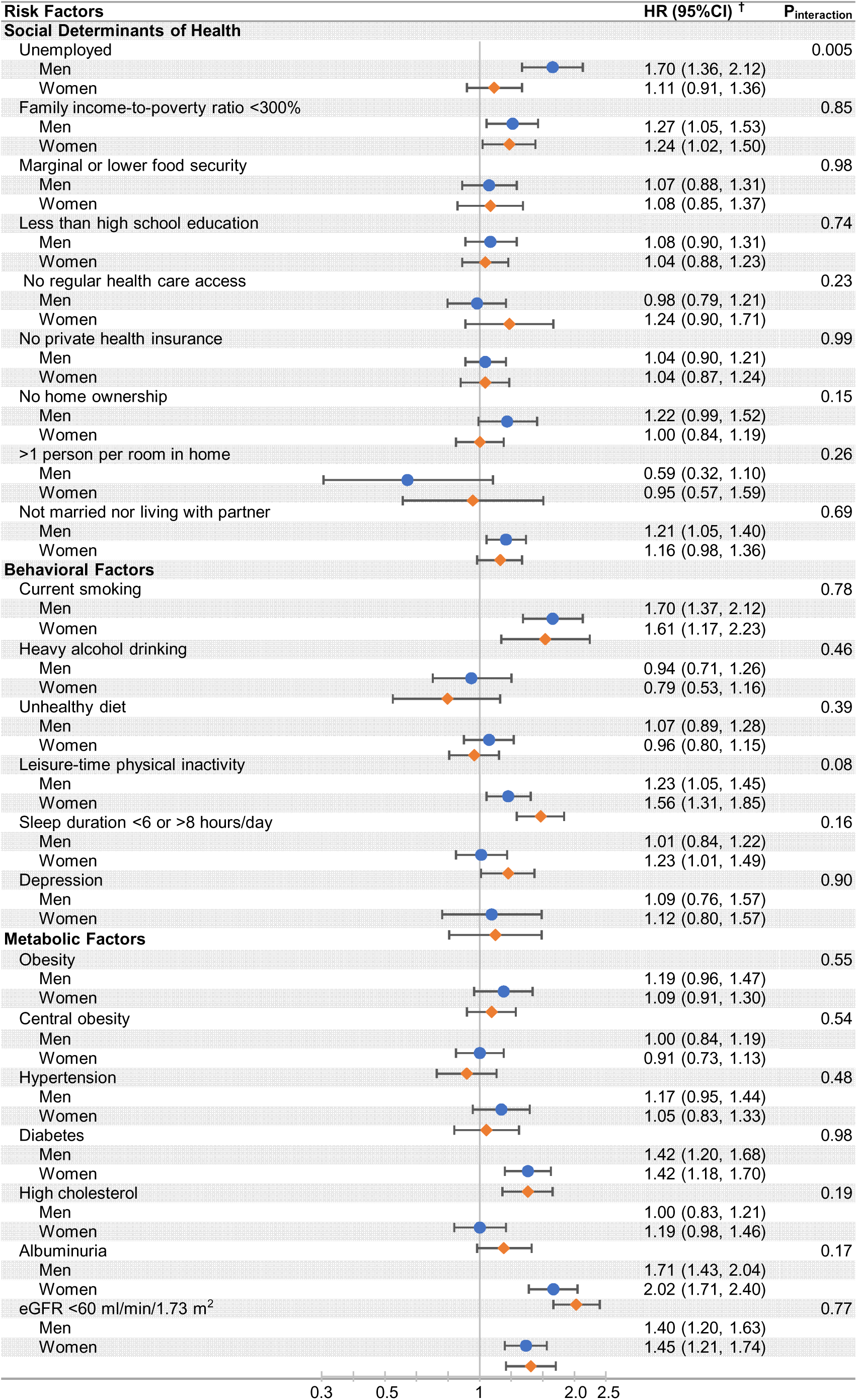
Multivariable-adjusted Associations of Risk Factors with Cardiovascular Disease Mortality by Sex Among Adults 60 Years of Age and Older *All numbers in the table are weighted to the US population and account for multiple imputation. Abbreviations: CI, confidence interval; HEI, healthy eating index; eGFR, estimated glomerular filtration rate. ^†^ Age is treated as the time scale and models are stratified by birth cohort and adjusted for sex, race, and all other SDOH, behavioral factors, and metabolic factors listed in the table.

In simultaneously considering all 22 risk factors, the average PAFs for CVD mortality was 67.9% (95% CI: 62.4-73.4) among men and 66.8% (95% CI: 60.6-73.1) among women **(Figure 5** and **Supplemental Table 6)**. Among men, hypertension (14.3%), albuminuria (10.0%), family income to poverty ratio <300% (8.7%), leisure-time physical inactivity (6.6%), and diabetes (6.3%) were the top US population-level contributors to CVD mortality **(Figure 5)**. Among women, leisure-time physical inactivity (14.7%), albuminuria (11.7%), family income to poverty ratio <300% (10.0%), eGFR <60 ml/min/1.73 m^2^ (8.9%), and diabetes (6.5%) were the top contributors. Similar patterns were observed across age groups (**Supplemental Table 6**).

**Figure 5.**
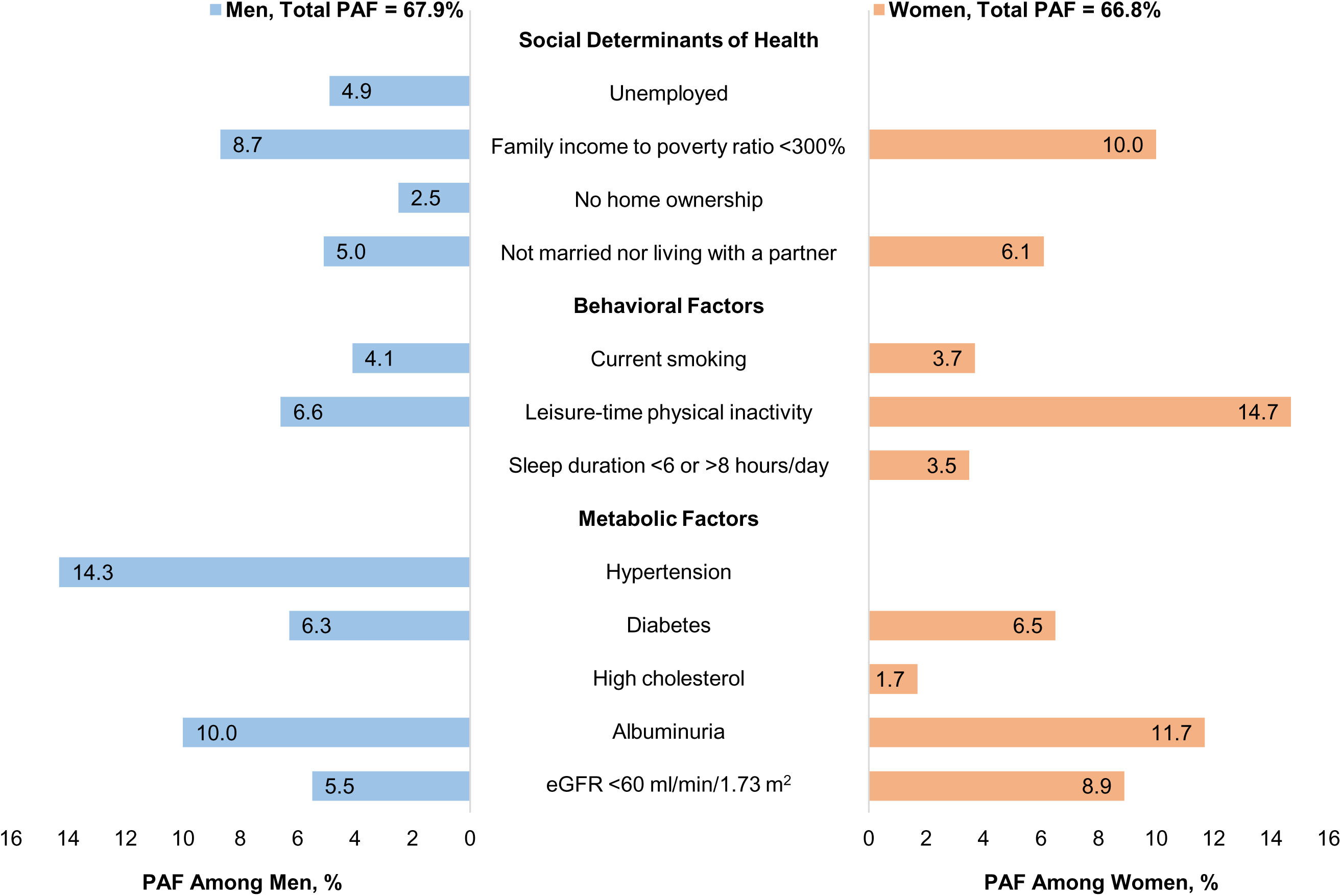
Population Attributable Fractions for Cardiovascular Disease Mortality Among US Adults by Sex *All numbers in the table are weighted to the US population and account for multiple imputation. All risk factors were included in one model and additionally adjusted for survey year, age, race/ethnicity, home ownership, education level, access to regular health care, room crowdedness, central obesity, depression score, ratio of total cholesterol and high-density lipoprotein cholesterol and body mass index. Abbreviations: eGFR, estimated glomerular filtration rate.

## Discussion

In this study including more than 50,000 adults from a representative sample of the US general population, we found that men had a more favorable SDOH profile, but less favorable behavioral and metabolic risk factor profile than women. Overall, men had higher absolute CVD mortality rates than women. However, the magnitudes of multivariable-adjusted associations between most risk factors and CVD mortality were similar among men and women. Therefore, sex differences in population-level CVD mortality (i.e., PAFs) were largely driven by sex differences in risk factor prevalence. The current findings suggest sex-specific efforts to prevent CVD mortality should focus on reducing the prevalence of risk factors particularly common among women or men.

Our results have important implications for primordial and primary prevention of CVD. Evaluation of sex differences in risk factors for CVD has gained considerable interest recently.^7,11,39,43–46^ However, previous studies have shown inconclusive results regarding whether associations of CVD risk factors with clinical outcomes differ between women and men.^4–6,12,39,46–49^ Our findings suggest that, among a generalizable sample of adults from the US general population, risk factor associations with CVD mortality are largely similar among men and women. Therefore, on a US population level, and for population CVD prevention, comprehensive strategies are needed to target these risk factors, and particularly upstream SDOH and behavioral factors. Incorporating SDOH risk factors into CVD risk assessment is essential.

As most recent guidelines state, SDOH risk factors play a critical role in the development of CVD events, independent of behavioral, lifestyle, and metabolic risk factors.^50–52^ In addition, the crucial role of behavioral and lifestyle risk factors in CVD risk is supported by several studies which reported that diet, exercise, alcohol consumption, and smoking together explained more than a half of overall PAFs among both men and women.^6,47^ Our results agree with these previous findings but extend them by showing they are independently associated with CVD mortality beyond a wide array of other risk factors.

Our study showed SDOH and behavioral collectively account for a third or more of the population-level burden of CVD mortality after adjusting for metabolic risk factors in both men and women. Few studies have evaluated sex differences in cardiovascular risk associated with SDOH, which are defined as the “structural determinants and conditions in which people are born, grow, live, work and age” that affect health, functioning, and quality of life.^19^ SDOH are strongly associated with premature death in the US general population, particularly from chronic causes like CVD and cancer.^53^ In the current study, compared with women, we found men were less likely to be unemployed, yet men who were unemployed had comparatively higher CVD mortality rates. The proportion of individuals taking care of the household or family among those unemployed was higher among women than men, which could partially explain these differences. Importantly, age played a significant role in some SDOH. For example, sex differences in the prevalence of family income-to-poverty ratio <300% and not being married nor living with a partner increased in older versus younger age groups. We also corroborate prior findings that many behavioral and lifestyle factors are strongly associated with CVD mortality in men and women, independent of SDOH and metabolic factors.^4–6,39^ However, we did not find sex differences in associations of current smoking, heavy alcohol drinking, unhealthy diet, and leisure-time physical inactivity with CVD mortality. These findings are different from previous studies,^4–6,39^ and may be attributable to differences in study design, study population, data collection, or analysis. Regardless, earlier screenings and lifestyle interventions were proposed recently to improve psychological health, which is supported by our results.^51^ Furthermore, policies and community strategies are needed to reduce SDOH burden in US adults, especially among young and middle age adults.

Since associations of risk factors with CVD mortality were mostly similar between men and women, the contribution of risk factors to PAFs varied between the sexes mainly due to the differences in prevalence of some risk factors. Of note, those with lower kidney function had the largest absolute CVD mortality rate compared with other risk factors, and lower kidney function was one of the top contributors to CVD mortality in both men and women regardless of age group. Additionally, top contributors for CVD mortality were hypertension and unemployment among men aged <60 years, diabetes among women aged <60 years, and leisure-time physical inactivity among both men and women aged ≥60 years. Therefore, our results support primordial prevention efforts targeting behavioral and lifestyle factors. Previous studies have suggested adherence to lifestyle guidelines involving these factors was associated with more than 80% lower risk of coronary heart disease among women.^6,54^

Most previous studies reported that associations between hypertension and diabetes with CVD events were stronger among women than men.^4–6,12,39^ We found the association of diabetes with CVD mortality was stronger among women than men before age 60 but similar after age 60.

Additionally, sex differences in prevalence of diabetes increased in old versus young and middle age group, with men older than 60 years having substantially higher prevalence of diabetes compared with women. Associations between hypertension and CVD mortality did not differ by sex. Furthermore, previous findings show that lipids increased greatly among women during the menopausal transition, but changes in insulin, glucose and BP were due more to chronological aging.^55^ This is in line with our finding that high cholesterol was associated with higher risk of CVD mortality among women than men when aged ≥60, but similar when aged <60. Men usually had a higher absolute risk than women for each risk factor across all age groups.

However, women had higher absolute risk than men in high cholesterol when aged ≥60 years. Prior reports suggest lipid control among women was worse than men.^10^ Of note, we observed that women had a worse kidney function profile which was associated with higher risk for CVD mortality compared with men. Since low kidney function is related to endothelial and microvascular dysfunction, potential reason for the difference could be that women experience more microvascular dysfunction than men.^56^

The current study has several strengths. First, the current analysis includes the most comprehensive simultaneous assessment of SDOH, behavioral factors, and metabolic factors among the US general population to date, with rigorous data collection methods across all cycles. Second, we evaluated the prevalence and magnitudes of association of 22 risk factors with CVD mortality, and combined them to estimate average PAFs to quantify their independent, population-level contributions to CVD mortality. However, the current study has several limitations. First, some important psychological metrics, like anxiety and stress, were unavailable. Furthermore, important sex-specific risk factors such as hormone status were unavailable owing to data limitations. However, since most women enter postmenopausal status by 60 years of age, we stratified analyses by age group which allows investigation of prevalence and associations pre- and post-menopause for most women.^5,6,39^ Second, secular trends in prevalence of risk factors and their associations with CVD mortality are possible with data collection spanning 20 years, although we accounted for birth cohort. Third, all risk factor data were collected simultaneously, limiting our ability to assess how risk factors may mediate each other. We aimed to present direct, independent associations of the risk factors with CVD mortality, but it should be acknowledged that the total associations of these risk factors with CVD mortality may be underestimated due to mediation. Fourth, participants aged <60 years had few CVD deaths, and thus statistical power in this age group is limited.

In conclusion, while the prevalence of SDOH, behavioral factors, and metabolic factors different among women compared with men, associations of these risk factors with CVD mortality were mostly similar by sex. SDOH and psycho-lifestyle risk factors collectively accounted for half of the average PAFs in both men and women. Low kidney function and leisure time physical inactivity were top contributors to CVD mortality in both men and women. For those less than 60 years of age, hypertension was top contributor to CVD mortality among men, and diabetes among women. Sex-specific prevention strategies targeting primordial prevention of risk factors should be developed among both men and women.

## Data Availability

All data produced are available online at: https://wwwn.cdc.gov/nchs/nhanes/Default.aspx

## Acknowledgments

The authors thank the NHANES study participants and study personnel for their time and effort. Portions of this manuscript were presented at the Annual Building Interdisciplinary Research Careers in Women’s Health (BIRCWH) Meeting, December 14, 2021, and the American Heart Association Epi/Lifestyle Scientific Sessions, March 2, 2023.

## Nonstandard Abbreviations and Acronyms

BMI: body mass index
BP: blood pressure
CI: confidence interval
CVD: cardiovascular disease
eGFR: estimated glomerular filtration rate
HbA1c: glycated hemoglobin
HEI-2015: Healthy Eating Index-2015
HR: hazard ratio
ICD-10: International Classification of Diseases, Tenth Revision
MEC: mobile examination center
NDI: National Death Index
NHANES: National Health and Nutrition Examination
Survey PHQ-9: Patient Health Questionnaire-9
SDOH: social determinants of health

## Supplemental Material

Tables S1–S6

