## Supplemental Tables 1-6 for "Sex Differences in Social, Behavioral, and Metabolic Risk Factors for Cardiovascular Disease Mortality among US Adults"

**Supplemental Materials**

Supplemental Table 1. Definitions of Social Determinants of Health, Behavioral Factors, and Metabolic Factors

Supplemental Table 2. Baseline Characteristics and Risk Factors Among US Men and Women by Age Group, NHANES 1999-2018

Supplemental Table 3. Multivariable-adjusted Associations of Risk Factors with Cardiovascular Disease Mortality by Sex Among All US Adults

Supplemental Table 4. Multivariable-adjusted Associations of Risk Factors with Cardiovascular Disease Mortality by Sex Among Adults Less than 60 Years of Age

Supplemental Table 5. Multivariable-adjusted Associations of Risk Factors with Cardiovascular Disease Mortality by Sex Among US Adults 60 Years of Age and Older

Supplemental Table 6. Population Attributable Fractions for Cardiovascular Diseases Mortality Among US Adults by Sex and Age Group

**Supplemental Table 1. Definitions of Social Determinants of Health, Behavioral Factors, and Metabolic Factors**

| **Risk Factors** | **Definitions** |
| --- | --- |
| **Social Determinants of Health** | |
| Employment status | Employed: currently working, or a student or retired; Not employed: taking care of house or family, unable to work for health reasons, on layoff, disabled, or other |
| Family income-to-poverty ratio | Divide family income by a poverty threshold related to family size and adjusted for geographic region and inflation |
| Food security | Full food security: 0 affirmative responses to questionnaire; Low or no food security: 1 or more affirmative responses |
| Education level | Greater than or equal to high school; Less than high school |
| Regular health care access | Routine place to go for healthcare; No routine place or ER/hospital/other |
| Health insurance status | Private insurance; Government or no health insurance |
| Home ownership | Own home: home owned or being bought; Not own home: home rented or other arrangement |
| Crowded housing | 1 person per room in the home or fewer; >1 person per room |
| Marital status | Married or living with a partner; Widowed/divorced/separated/never married |
| **Behavioral Factors** | |
| Current smoking | Formerly or never smoke; Currently smoke |
| Heavy alcohol drinking | >14 drinks per week for men, and >7 for women |
| Low diet quality | High diet quality: Healthy Eating Index-2015 ≥52 (median); Healthy Eating Index-2015 <52 |
| Leisure-time physical inactivity | Have leisure-time physical activity; No leisure-time physical activity |
| Sleep duration | Normal: 6< sleep <8 hours/day; Excessive/deprived sleep: sleep >8 or <6 hours/day |
| Depression | Patient Health Questionnaire-9 <10; Patient Health Questionnaire-9 ≥10 |
| **Metabolic Factors** | |
| Obesity | Body mass index ≥30 kg/m^2^ |
| Central obesity | Waist circumference ≥102 cm for men and ≥88 cm for women |
| Hypertension | No hypertension: systolic blood pressure <130 mm Hg and diastolic blood pressure <80 mm Hg and no antihypertensive medication  Hypertension: systolic blood pressure ≥130 mm Hg or diastolic blood pressure ≥80 mm Hg or antihypertensive medication |
| Diabetes | Fasting glucose ≥126 mg/dL or hemoglobin A1c ≥6.5%, or diagnosed diabetes |
| High cholesterol | Ratio of total to HDL cholesterol ≥5 |
| Urinary albumin-to-creatinine ratio | Urinary albumin-to-creatinine ratio <30 mg/g |
| Estimated glomerular filtration rate | Estimated glomerular filtration rate <60 ml/min/1.73m^2^ (CKD-EPI Creatinine-based Equation 2021) |

**Supplemental Table 2. Baseline Characteristics and Risk Factors Among US Men and Women by Age Group, NHANES 1999-2018**

| **Characteristics ^*^** | **Age <60 years (N = 33,023)** | | | **Age ≥60 years (N = 17,785)** | | |
| --- | --- | --- | --- | --- | --- | --- |
|  | **Men (N = 16,359)** | **Women (N = 16,664)** | **P-value** | **Men (N = 8,784)** | **Women (N = 9,001)** | **P-value** |
| **Social determinants of health, %** ^†^ | | | | | | |
| Unemployed | 14.0 (13.2, 14.8) | 27.3 (26.2, 28.3) | <0.001 | 10.9 (9.8, 11.9) | 17.7 (16.5, 18.9) | <0.001 |
| Family income-to-poverty ratio <300% | 48.6 (47.1, 50.2) | 51.6 (50.0, 53.2) | <0.001 | 48.5 (46.5, 50.6) | 58.5 (56.2, 60.7) | <0.001 |
| Marginal or lower food security | 23.0 (22.0, 24.1) | 25.0 (23.8, 26.1) | <0.001 | 11.7 (10.8, 12.7) | 14.2 (13.0, 15.4) | <0.001 |
| Less than high school education | 17.1 (16.1, 18.1) | 14.8 (13.9, 15.7) | <0.001 | 21.6 (20.1, 23.2) | 22.9 (21.4, 24.4) | 0.08 |
| No regular health care access | 31.6 (30.5, 32.7) | 16.6 (15.8, 17.5) | <0.001 | 10.3 (9.5, 11.1) | 6.3 (5.7, 6.9) | <0.001 |
| No private health insurance | 36.2 (34.9, 37.5) | 34.7 (33.4, 36.1) | 0.009 | 39.7 (38.0, 41.4) | 40.7 (39.1, 42.4) | 0.18 |
| No home ownership | 37.6 (36.1, 39.1) | 36.6 (35.1, 38.1) | 0.03 | 15.2 (14.0, 16.5) | 19.4 (17.7, 21.0) | <0.001 |
| >1 person per room in home | 7.4 (6.7, 8.2) | 6.8 (6.1, 7.4) | 0.01 | 1.6 (1.3, 1.9) | 1.6 (1.2, 1.9) | 0.82 |
| Not married nor living with a partner | 35.8 (34.6, 37.0) | 37.3 (36.2, 38.5) | 0.007 | 23.1 (21.7, 24.5) | 49.2 (47.6, 50.8) | <0.001 |
| **Behavioral risk factors, %** ^‡^ | | | | | | |
| Current smoking | 27.7 (26.6, 28.8) | 22.6 (21.6, 23.6) | <0.001 | 13.7 (12.6, 14.7) | 9.9 (9.1, 10.7) | <0.001 |
| Heavy alcohol drinking | 12.6 (12.0, 13.3) | 8.7 (8.0, 9.4) | <0.001 | 10.0 (8.9, 11.0) | 7.0 (6.0, 7.9) | <0.001 |
| Unhealthy diet | 57.8 (56.5, 59.1) | 51.0 (49.4, 52.5) | <0.001 | 41.5 (39.9, 43.1) | 33.5 (31.9, 35.1) | <0.001 |
| Leisure-time physical inactivity | 37.4 (36.0, 38.8) | 40.5 (39.2, 41.7) | <0.001 | 50.2 (48.4, 51.9) | 58.1 (56.2, 60.0) | <0.001 |
| Sleep duration <6 or >8 hours/day | 22.3 (21.3, 23.2) | 24.7 (23.7, 25.6) | <0.001 | 26.5 (25.2, 27.9) | 29.0 (27.7, 30.4) | 0.006 |
| Depression | 6.3 (5.8, 6.8) | 9.9 (9.2, 10.5) | <0.001 | 5.3 (4.5, 6.1) | 7.7 (6.9, 8.5) | <0.001 |
| **Metabolic risk factors, % ^§^** | | | | | | |
| Obesity | 33.4 (32.2, 34.6) | 36.6 (35.5, 37.7) | <0.001 | 35.3 (33.7, 36.9) | 37.7 (36.2, 39.2) | 0.02 |
| Central obesity | 40.2 (38.9, 41.5) | 59.6 (58.4, 60.8) | <0.001 | 59.0 (57.4, 60.6) | 76.2 (74.8, 77.6) | <0.001 |
| Hypertension | 42.4 (41.2, 43.6) | 31.6 (30.6, 32.6) | <0.001 | 74.8 (73.2, 76.4) | 78.2 (77.0, 79.5) | <0.001 |
| Diabetes | 9.0 (8.3, 9.7) | 7.8 (7.3, 8.4) | 0.006 | 29.9 (28.4, 31.4) | 23.3 (22.0, 24.6) | <0.001 |
| High cholesterol | 30.8 (29.8, 31.8) | 12.7 (12.0, 13.4) | <0.001 | 20.5 (19.1, 21.8) | 13.2 (12.3, 14.2) | <0.001 |
| Albuminuria | 6.2 (5.7, 6.7) | 8.1 (7.6, 8.6) | <0.001 | 19.7 (18.6, 20.8) | 17.5 (16.3, 18.7) | 0.006 |
| eGFR <60 ml/min/1.73 m^2^ | 1.1 (0.9, 1.3) | 1.4 (1.1, 1.6) | 0.11 | 17.2 (16.1, 18.2) | 22.2 (21.1, 23.2) | <0.001 |

Abbreviations: CI, confidence interval; eGFR, estimated glomerular filtration rate; NHANES, National Health and Nutrition Examination Survey.

^*^ Sample sizes are unobserved frequencies while the other numbers in the table are weighted means (95% CI) or percentages (95% CI) after multiple imputation.

^†^ Employment is defined as currently working, student, or retired. Food insecurity is assessed with the 10-item adult US Food Security Survey Module with zero affirmative responses indicating high food security and ≥1 affirmative responses indicating marginal or lower food security.

^‡^ Heavy alcohol drinking is defined with more than 14 drinks per week for men, and 7 for women. The Healthy Eating Index (HEI) uses a scoring system to assess how well a set of foods aligns with key recommendations of the Dietary Guidelines for Americans 2015-2020. The scores range from 0 to 100 with a higher score indicating greater consistency of the diet with the Dietary Guidelines for Americans. Median HEI score was 52 among NHANES 1999-2018 participants; Depression was assessed with the Patient Health Questionnaire-9 (PHQ-9) with scores ≥10 indicating depression.

**^§^** Obesity is defined as body mass index ≥30 kg/m^2^; central obesity is defined as waist circumference ≥102 cm for men and ≥88 cm for women; hypertension is defined as systolic blood pressure ≥130 mmHg and/or diastolic blood pressure ≥80 mmHg or use of antihypertensive medications; and diabetes is defined as fasting glucose ≥126mg/dL or hemoglobin A1c ≥6.5% or diagnosed diabetes.

**Supplemental Table 3. Multivariable-adjusted Associations of Risk Factors with Cardiovascular Disease Mortality by Sex Among All US Adults ^*^**

| **Risk factors** | **CVD Mortality / # of Participants, (%)** | | **Minimally Adjusted ^\|\|^** | | | **Fully Adjusted ^#^** | | |
| --- | --- | --- | --- | --- | --- | --- | --- | --- |
|  | **Men** | **Women** | **Men** | **Women** | **P-value** | **Men** | **Women** | **P-value** |
| **Social determinants of health ^†^** | | | | | | | | |
| Employment status |  |  |  |  |  |  |  |  |
| Employed, student, or retired | 1,170/20,938 (5.6) | 859/18,336 (4.7) | Ref | Ref | <0.001 | Ref | Ref | <0.001 |
| Unemployed | 279/4,189 (6.7) | 281/7,307 (3.8) | 3.03 (2.54, 3.61) | 1.53 (1.29, 1.81) |  | 1.97 (1.62, 2.39) | 1.19 (1.00, 1.43) |  |
| Family income-to-poverty ratio |  |  |  |  |  |  |  |  |
| ≥300% | 380/8,884 (4.3) | 219/8,212 (2.7) | Ref | Ref | 0.68 | Ref | Ref | 0.88 |
| <300% | 959/14,063 (6.8) | 781/15,058 (5.2) | 1.82 (1.55, 2.15) | 1.92 (1.61, 2.28) |  | 1.25 (1.05, 1.50) | 1.28 (1.06, 1.54) |  |
| Food security |  |  |  |  |  |  |  |  |
| Full security | 1,139/17,949 (6.3) | 895/17,760 (5.0) | Ref | Ref | 0.32 | Ref | Ref | 0.80 |
| Marginal, low, or very low security | 280/6,605 (4.2) | 212/7,316 (2.9) | 1.97 (1.62, 2.39) | 1.72 (1.40, 2.10) |  | 1.15 (0.95, 1.40) | 1.11 (0.87, 1.40) |  |
| Education level |  |  |  |  |  |  |  |  |
| High school graduate or higher | 849/17,980 (4.7) | 633/18,852 (3.4) | Ref | Ref | 0.87 | Ref | Ref | 0.65 |
| Less than high school | 594/7,127 (8.3) | 500/6,768 (7.4) | 1.55 (1.30, 1.83) | 1.52 (1.31, 1.76) |  | 1.11 (0.92, 1.33) | 1.04 (0.89, 1.22) |  |
| Regular health care access |  |  |  |  |  |  |  |  |
| At least one regular health care facility | 1,265/18,105 (7.0) | 1,044/21,653 (4.8) | Ref | Ref | 0.37 | Ref | Ref | 0.21 |
| None or emergency room | 184/7,035 (2.6) | 95/4,010 (2.4) | 1.08 (0.86, 1.35) | 1.27 (0.99, 1.63) |  | 0.92 (0.73, 1.16) | 1.18 (0.89, 1.57) |  |
| Health insurance status |  |  |  |  |  |  |  |  |
| Private | 698/13,179 (5.3) | 514/13,586 (3.8) | Ref | Ref | 0.70 | Ref | Ref | 0.71 |
| Government or none | 731/11,695 (6.3) | 605/11,816 (5.1) | 1.48 (1.28, 1.72) | 1.42 (1.21, 1.68) |  | 1.03 (0.89, 1.20) | 1.08 (0.90, 1.29) |  |
| Home ownership |  |  |  |  |  |  |  |  |
| Own home | 1,028/15,573 (6.6) | 761/15,724 (4.8) | Ref | Ref | 0.09 | Ref | Ref | 0.24 |
| Rent home or other arrangement | 405/9,169 (4.4) | 358/9,547 (3.7) | 1.77 (1.52, 2.07) | 1.48 (1.28, 1.70) |  | 1.24 (1.03, 1.48) | 1.07 (0.91, 1.25) |  |
| Number of persons per room in home |  |  |  |  |  |  |  |  |
| ≤1 person | 1,386/22,460 (6.2) | 1,069/23,022 (4.6) | Ref | Ref | 0.51 | Ref | Ref | 0.46 |
| >1 person | 46/2,231 (2.1) | 45/2,222 (2.0) | 1.11 (0.70, 1.76) | 1.33 (0.89, 1.99) |  | 0.86 (0.54, 1.38) | 1.08 (0.70, 1.66) |  |
| Marital status |  |  |  |  |  |  |  |  |
| Married or living with a partner | 908/16,320 (5.6) | 354/13,618 (2.6) | Ref | Ref | 0.40 | Ref | Ref | 0.35 |
| Not married nor living with a partner | 522/8,599 (6.1) | 758/11,803 (6.4) | 1.72 (1.49, 1.98) | 1.57 (1.34, 1.84) |  | 1.30 (1.11, 1.52) | 1.17 (0.99, 1.38) |  |
| **Behavioral factors ^‡^** | | | | | | | | |
| Smoking |  |  |  |  |  |  |  |  |
| Former or never smoking | 1,157/18,857 (6.1) | 982/21,120 (4.6) | Ref | Ref | 0.59 | Ref | Ref | 0.49 |
| Current smoking | 290/6,260 (4.6) | 154/4,520 (3.4) | 2.16 (1.83, 2.55) | 2.02 (1.65, 2.46) |  | 1.66 (1.38, 2.00) | 1.84 (1.47, 2.30) |  |
| Alcohol drinking |  |  |  |  |  |  |  |  |
| Moderate or no alcohol drinking | 1,223/20,698 (5.9) | 984/21,461 (4.6) | Ref | Ref | 0.02 | Ref | Ref | 0.22 |
| Heavy alcohol drinking | 121/2,449 (4.9) | 48/1,512 (3.2) | 1.12 (0.89, 1.40) | 0.65 (0.44, 0.94) |  | 1.07 (0.85, 1.35) | 0.81 (0.54, 1.20) |  |
| Diet quality |  |  |  |  |  |  |  |  |
| HEI ≥52 | 640/10,070 (6.4) | 565/11,956 (4.7) | Ref | Ref | 0.44 | Ref | Ref | 0.31 |
| HEI <52 | 594/10,786 (5.5) | 401/9,837 (4.1) | 1.39 (1.17, 1.65) | 1.27 (1.09, 1.49) |  | 1.15 (0.97, 1.36) | 1.02 (0.87, 1.20) |  |
| Leisure-time physical activity |  |  |  |  |  |  |  |  |
| Any | 547/13,239 (4.1) | 295/12,086 (2.4) | Ref | Ref | 0.28 | Ref | Ref | 0.15 |
| No | 900/11,871 (7.6) | 844/13,559 (6.2) | 1.71 (1.49, 1.96) | 1.94 (1.64, 2.29) |  | 1.20 (1.03, 1.40) | 1.44 (1.21, 1.72) |  |
| Sleep duration |  |  |  |  |  |  |  |  |
| 6-8 hours per day | 488/13,438 (3.6) | 339/13,358 (2.5) | Ref | Ref | 0.48 | Ref | Ref | 0.52 |
| <6 or >8 hours per day | 224/4,921 (4.6) | 204/5,555 (3.7) | 1.20 (0.99, 1.45) | 1.32 (1.11, 1.58) |  | 1.08 (0.90, 1.29) | 1.17 (0.97, 1.42) |  |
| Depression |  |  |  |  |  |  |  |  |
| PHQ-9 <10 | 585/15,596 (3.8) | 386/14,868 (2.6) | Ref | Ref | 0.98 | Ref | Ref | 0.45 |
| PHQ-9 ≥10 | 49/1,101 (4.5) | 65/1,864 (3.5) | 1.65 (1.16, 2.33) | 1.66 (1.24, 2.20) |  | 1.05 (0.73, 1.49) | 1.23 (0.90, 1.68) |  |
| **Metabolic factors ^§^** | | | | | | | | |
| Obesity |  |  |  |  |  |  |  |  |
| No | 930/16,584 (5.6) | 629/15,074 (4.2) | Ref | Ref | 0.31 | Ref | Ref | 0.71 |
| Yes | 418/8,037 (5.2) | 420/10,078 (4.2) | 1.49 (1.30, 1.72) | 1.35 (1.19, 1.54) |  | 1.18 (0.98, 1.42) | 1.13 (0.96, 1.32) |  |
| Central obesity |  |  |  |  |  |  |  |  |
| No | 593/13,511 (4.4) | 229/7,806 (2.9) | Ref | Ref | 0.29 | Ref | Ref | 0.23 |
| Yes | 703/10,314 (6.8) | 732/16,246 (4.5) | 1.35 (1.19, 1.53) | 1.22 (1.05, 1.41) |  | 1.04 (0.88, 1.23) | 0.89 (0.73, 1.08) |  |
| Hypertension |  |  |  |  |  |  |  |  |
| No | 256/10,799 (2.4) | 144/12,216 (1.2) | Ref | Ref | 0.67 | Ref | Ref | 0.62 |
| Yes | 1,148/13,510 (8.5) | 962/12,439 (7.7) | 1.68 (1.41, 2.00) | 1.77 (1.42, 2.21) |  | 1.43 (1.19, 1.72) | 1.33 (1.03, 1.70) |  |
| Diabetes |  |  |  |  |  |  |  |  |
| No | 973/20,849 (4.7) | 778/21,662 (3.6) | Ref | Ref | 0.75 | Ref | Ref | 0.61 |
| Yes | 476/4,294 (11.1) | 362/4,002 (9.0) | 2.03 (1.73, 2.38) | 2.11 (1.82, 2.44) |  | 1.45 (1.22, 1.73) | 1.55 (1.31, 1.84) |  |
| High cholesterol |  |  |  |  |  |  |  |  |
| No | 1,001/17,118 (5.8) | 846/20,742 (4.1) | Ref | Ref | 0.05 | Ref | Ref | 0.16 |
| Yes | 344/6,532 (5.3) | 197/3,294 (6.0) | 1.21 (1.05, 1.40) | 1.54 (1.29, 1.84) |  | 1.06 (0.91, 1.24) | 1.26 (1.05, 1.52) |  |
| Albumin-to-creatinine ratio |  |  |  |  |  |  |  |  |
| <30 mg/g | 900/21,318 (4.2) | 674/21,691 (3.1) | Ref | Ref | 0.45 | Ref | Ref | 0.56 |
| ≥30 mg/g | 475/3,186 (14.9) | 368/3,254 (11.3) | 2.60 (2.23, 3.03) | 2.81 (2.38, 3.33) |  | 1.94 (1.65, 2.28) | 2.08 (1.75, 2.47) |  |
| Estimated glomerular filtration rate |  |  |  |  |  |  |  |  |
| ≥60 ml/min/1.73 m^2^ | 957/21,702 (4.4) | 639/21,820 (2.9) | Ref | Ref | 0.44 | Ref | Ref | 0.57 |
| <60 ml/min/1.73 m^2^ | 389/1,900 (20.5) | 401/2,145 (18.7) | 1.75 (1.49, 2.06) | 1.91 (1.63, 2.24) |  | 1.40 (1.18, 1.66) | 1.51 (1.26, 1.79) |  |

Abbreviations: CI, confidence interval; HEI, healthy eating index; PHQ-9, Patient Health Questionnaire-9; NHANES, National Health and Nutrition Examination Survey.

^*^ CVD mortality/number of participants is unweighted using original data. All the other numbers in the table are weighted accounting for the complex, multistage, probability design of NHANES and multiple imputation. For Cox regression, age is treated as timescale and models are stratified by birth cohort.

^†^ Employment is defined as at work, student or retired. Food insecurity is assessed with the 18-item US Food Security Survey Module with zero affirmative responses indicating high food security and ≥1 affirmative responses indicating marginal or lower food security.

^‡^ Heavy alcohol drinking is defined with more than 14 (7) drinks per week for men (women); The Healthy Eating Index (HEI) uses a scoring system to assess how well a set of foods aligns with key recommendations of the Dietary Guidelines for Americans. The scores range from 0 to 100 with a higher score indicating greater consistency of the diet with the Dietary Guidelines for Americans. Median HEI score was 52 among NHANES 1999-2018 participants; Depression was assessed with the Patient Health Questionnaire-9 (PHQ-9) with scores ≥10 indicating depression.

**^§^** Obesity is defined as body mass index ≥30 kg/m^2^; central obesity is defined as waist circumference ≥102 cm for men and ≥88 cm for women; hypertension is defined as systolic blood pressure ≥130 mmHg and/or diastolic blood pressure ≥80 mmHg or use of antihypertensive medications; and diabetes is defined as fasting glucose ≥126mg/dL or hemoglobin A1c ≥6.5% or diagnosed diabetes.

**^||^** Adjusted for sex and race. For Cox regression, age is treated as the timescale and models are stratified by birth cohort.

**^#^** Adjusted for sex, race, and all other psychosocial, behavioral, and metabolic risk factors listed in the table. For Cox regression, age is treated as timescale and models are stratified by birth cohort.

**Supplemental Table 4. Multivariable-adjusted Associations of Risk Factors with Cardiovascular Disease Mortality by Sex Among Adults Less than 60 Years of Age ^*^**

| **Risk factors** | **CVD Mortality / # of Participants, (%)** | | **Minimally Adjusted ^\|\|^** | | | **Fully Adjusted ^#^** | | |
| --- | --- | --- | --- | --- | --- | --- | --- | --- |
|  | **Men** | **Women** | **Men** | **Women** | **P-value** | **Men** | **Women** | **P-value** |
| **Social determinants of health ^†^** | | | | | | | | |
| Employment status |  |  |  |  |  |  |  |  |
| Employed, student, or retired | 128/13,423 (1.0) | 64/11,414 (0.6) | Ref | Ref | 0.10 | Ref | Ref | 0.04 |
| Unemployed | 105/2,925 (3.6) | 76/5,236 (1.5) | 3.99 (2.81, 5.66) | 2.53 (1.71, 3.74) |  | 2.23 (1.50, 3.30) | 1.19 (0.81, 1.74) |  |
| Family income-to-poverty ratio |  |  |  |  |  |  |  |  |
| ≥300% | 56/5,984 (0.9) | 26/5,715 (0.5) | Ref | Ref | 0.34 | Ref | Ref | 0.66 |
| <300% | 155/9,034 (1.7) | 99/9,582 (1.0) | 2.55 (1.75, 3.72) | 3.42 (2.12, 5.49) |  | 1.13 (0.70, 1.82) | 1.34 (0.74, 2.45) |  |
| Food security |  |  |  |  |  |  |  |  |
| Full security | 138/10,999 (1.3) | 80/10,853 (0.7) | Ref | Ref | 0.95 | Ref | Ref | 0.55 |
| Marginal, low, or very low security | 89/4,959 (1.8) | 54/5,430 (1.0) | 2.56 (1.77, 3.70) | 2.61 (1.71, 3.97) |  | 1.29 (0.85, 1.95) | 1.05 (0.64, 1.72) |  |
| Education level |  |  |  |  |  |  |  |  |
| High school graduate or higher | 144/12,265 (1.2) | 83/12,998 (0.6) | Ref | Ref | 0.30 | Ref | Ref | 0.70 |
| Less than high school | 88/4,081 (2.2) | 57/3,653 (1.6) | 2.00 (1.36, 2.95) | 2.76 (1.76, 4.34) |  | 1.20 (0.82, 1.74) | 1.36 (0.84, 2.20) |  |
| Regular health care access |  |  |  |  |  |  |  |  |
| At least one regular health care facility | 177/10,482 (1.7) | 115/13,402 (0.9) | Ref | Ref | 0.46 | Ref | Ref | 0.44 |
| None or emergency room | 56/5,875 (1.0) | 25/3,261 (0.8) | 1.02 (0.69, 1.52) | 1.29 (0.79, 2.12) |  | 0.85 (0.55, 1.30) | 1.11 (0.65, 1.89) |  |
| Health insurance status |  |  |  |  |  |  |  |  |
| Private | 106/8,761 (1.2) | 52/9,153 (0.6) | Ref | Ref | 0.29 | Ref | Ref | 0.32 |
| Government or none | 123/7,393 (1.7) | 84/7,320 (1.1) | 2.46 (1.75, 3.47) | 3.21 (2.10, 4.90) |  | 0.96 (0.63, 1.47) | 1.33 (0.81, 2.17) |  |
| Home ownership |  |  |  |  |  |  |  |  |
| Own home | 116/8,874 (1.3) | 66/9,164 (0.7) | Ref | Ref | 0.51 | Ref | Ref | 0.92 |
| Rent home or other arrangement | 114/7,214 (1.6) | 71/7,252 (1.0) | 2.11 (1.52, 2.92) | 2.48 (1.76, 3.48) |  | 1.27 (0.88, 1.83) | 1.23 (0.84, 1.80) |  |
| Number of persons per room in home |  |  |  |  |  |  |  |  |
| ≤1 person | 205/14,148 (1.4) | 121/14,518 (0.8) | Ref | Ref | 0.59 | Ref | Ref | 0.64 |
| >1 person | 25/1,913 (1.3) | 16/1,888 (0.8) | 1.30 (0.69, 2.42) | 1.63 (0.86, 3.08) |  | 1.07 (0.56, 2.04) | 1.31 (0.68, 2.51) |  |
| Marital status |  |  |  |  |  |  |  |  |
| Married or living with a partner | 120/10,115 (1.2) | 54/9,684 (0.6) | Ref | Ref | 0.66 | Ref | Ref | 0.49 |
| Not married nor living with a partner | 106/6,076 (1.7) | 82/6,844 (1.2) | 2.37 (1.65, 3.40) | 2.11 (1.34, 3.33) |  | 1.65 (1.09, 2.50) | 1.34 (0.80, 2.24) |  |
| **Behavioral factors ^‡^** | | | | | | | | |
| Smoking |  |  |  |  |  |  |  |  |
| Former or never smoking | 126/11,461 (1.1) | 75/13,022 (0.6) | Ref | Ref | 0.43 | Ref | Ref | 0.45 |
| Current smoking | 107/4,885 (2.2) | 64/3,631 (1.8) | 2.29 (1.68, 3.12) | 2.89 (1.93, 4.33) |  | 1.58 (1.08, 2.31) | 2.04 (1.29, 3.23) |  |
| Alcohol drinking |  |  |  |  |  |  |  |  |
| Moderate or no alcohol drinking | 175/13,202 (1.3) | 121/13,607 (0.9) | Ref | Ref | 0.16 | Ref | Ref | 0.25 |
| Heavy alcohol drinking | 36/1,754 (2.1) | 8/1,083 (0.7) | 1.46 (0.98, 2.18) | 0.76 (0.34, 1.73) |  | 1.33 (0.87, 2.03) | 0.76 (0.32, 1.78) |  |
| Diet quality |  |  |  |  |  |  |  |  |
| HEI ≥52 | 63/5,773 (1.1) | 44/6,929 (0.6) | Ref | Ref | 0.86 | Ref | Ref | 0.98 |
| HEI <52 | 132/7,629 (1.7) | 80/7,159 (1.1) | 1.83 (1.28, 2.61) | 1.93 (1.20, 3.12) |  | 1.40 (0.96, 2.03) | 1.41 (0.86, 2.30) |  |
| Leisure-time physical activity |  |  |  |  |  |  |  |  |
| Any | 96/9,465 (1.0) | 48/8,820 (0.5) | Ref | Ref | 0.99 | Ref | Ref | 0.73 |
| No | 136/6,872 (2.0) | 92/7,832 (1.2) | 1.90 (1.35, 2.69) | 1.90 (1.25, 2.88) |  | 1.16 (0.82, 1.65) | 1.04 (0.65, 1.66) |  |
| Sleep duration |  |  |  |  |  |  |  |  |
| 6-8 hours per day | 58/9,055 (0.6) | 43/9,032 (0.5) | Ref | Ref | 0.52 | Ref | Ref | 0.36 |
| <6 or >8 hours per day | 42/3,000 (1.4) | 24/3,430 (0.7) | 1.55 (0.99, 2.41) | 1.22 (0.69, 2.17) |  | 1.27 (0.82, 1.98) | 0.88 (0.47, 1.65) |  |
| Depression |  |  |  |  |  |  |  |  |
| PHQ-9 <10 | 74/10,199 (0.7) | 41/9,613 (0.4) | Ref | Ref | 0.30 | Ref | Ref | 0.21 |
| PHQ-9 ≥10 | 14/749 (1.9) | 22/1,313 (1.7) | 1.92 (1.11, 3.32) | 2.80 (1.57, 4.99) |  | 0.87 (0.46, 1.64) | 1.46 (0.78, 2.73) |  |
| **Metabolic factors ^§^** | | | | | | | | |
| Obesity |  |  |  |  |  |  |  |  |
| No | 139/10,842 (1.3) | 57/9,857 (0.6) | Ref | Ref | 0.59 | Ref | Ref | 0.93 |
| Yes | 82/5,308 (1.5) | 77/6,591 (1.2) | 1.70 (1.23, 2.35) | 1.97 (1.30, 2.96) |  | 1.09 (0.68, 1.74) | 1.13 (0.63, 2.00) |  |
| Central obesity |  |  |  |  |  |  |  |  |
| No | 108/9,794 (1.1) | 27/5,960 (0.5) | Ref | Ref | 0.96 | Ref | Ref | 0.39 |
| Yes | 104/5,912 (1.8) | 102/9,935 (1.0) | 1.73 (1.25, 2.41) | 1.76 (1.10, 2.81) |  | 1.14 (0.72, 1.79) | 0.82 (0.44, 1.53) |  |
| Hypertension |  |  |  |  |  |  |  |  |
| No | 53/8,882 (0.6) | 35/10,614 (0.3) | Ref | Ref | 0.58 | Ref | Ref | 0.93 |
| Yes | 170/6,837 (2.5) | 97/5,240 (1.9) | 2.68 (1.88, 3.83) | 3.18 (1.96, 5.16) |  | 2.02 (1.35, 3.03) | 2.09 (1.23, 3.54) |  |
| Diabetes |  |  |  |  |  |  |  |  |
| No | 166/14,760 (1.1) | 84/15,119 (0.6) | Ref | Ref | 0.06 | Ref | Ref | 0.08 |
| Yes | 67/1,599 (4.2) | 56/1,545 (3.6) | 3.08 (2.07, 4.60) | 5.38 (3.40, 8.52) |  | 1.59 (1.00, 2.53) | 2.81 (1.69, 4.65) |  |
| High cholesterol |  |  |  |  |  |  |  |  |
| No | 120/10,616 (1.1) | 94/13,588 (0.7) | Ref | Ref | 0.04 | Ref | Ref | 0.37 |
| Yes | 90/4,752 (1.9) | 33/2,093 (1.6) | 1.43 (1.02, 2.01) | 2.48 (1.54, 3.99) |  | 1.13 (0.79, 1.63) | 1.45 (0.89, 2.35) |  |
| Albumin-to-creatinine ratio |  |  |  |  |  |  |  |  |
| <30 mg/g | 158/14,849 (1.1) | 89/14,838 (0.6) | Ref | Ref | 0.61 | Ref | Ref | 0.38 |
| ≥30 mg/g | 68/1,172 (5.8) | 42/1,517 (2.8) | 5.43 (3.95, 7.47) | 4.71 (2.92, 7.60) |  | 2.95 (2.00, 4.33) | 2.20 (1.34, 3.63) |  |
| Estimated glomerular filtration rate |  |  |  |  |  |  |  |  |
| ≥60 ml/min/1.73 m^2^ | 194/15,130 (1.3) | 106/15,404 (0.7) | Ref | Ref | 0.09 | Ref | Ref | 0.06 |
| <60 ml/min/1.73 m^2^ | 18/213 (8.5) | 20/231 (8.7) | 3.98 (2.08, 7.60) | 7.54 (3.95, 14.40) |  | 2.10 (1.08, 4.11) | 4.52 (2.44, 8.37) |  |

Abbreviations: CI, confidence interval; HEI, healthy eating index; PHQ-9, Patient Health Questionnaire-9; NHANES, National Health and Nutrition Examination Survey.

^*^ CVD mortality/number of participants is unweighted using original data. All the other numbers in the table are weighted accounting for the complex, multistage, probability design of NHANES and multiple imputation. For Cox regression, age is treated as timescale and models are stratified by birth cohort.

^†^ Employment is defined as at work, student or retired. Food insecurity is assessed with the 18-item US Food Security Survey Module with zero affirmative responses indicating high food security and ≥1 affirmative responses indicating marginal or lower food security.

^‡^ Heavy alcohol drinking is defined with more than 14 (7) drinks per week for men (women); The Healthy Eating Index (HEI) uses a scoring system to assess how well a set of foods aligns with key recommendations of the Dietary Guidelines for Americans. The scores range from 0 to 100 with a higher score indicating greater consistency of the diet with the Dietary Guidelines for Americans. Median HEI score was 52 among NHANES 1999-2018 participants; Depression was assessed with the Patient Health Questionnaire-9 (PHQ-9) with scores ≥10 indicating depression.

^§^ Obesity is defined as body mass index ≥30 kg/m^2^; central obesity is defined as waist circumference ≥102 cm for men and ≥88 cm for women; hypertension is defined as systolic blood pressure ≥130 mmHg and/or diastolic blood pressure ≥80 mmHg or use of antihypertensive medications; and diabetes is defined as fasting glucose ≥126mg/dL or hemoglobin A1c ≥6.5% or diagnosed diabetes.

**^||^** Adjusted for sex and race. For Cox regression, age is treated as timescale and models are stratified by birth cohort.

**^#^** Adjusted for sex, race, and all other psychosocial, behavioral, and metabolic risk factors listed in the table. For Cox regression, age is treated as timescale and models are stratified by birth cohort.

**Supplemental Table 5. Multivariable-adjusted Associations of Risk Factors with Cardiovascular Disease Mortality by Sex Among Adults 60 Years of Age and Older ^*^**

| **Risk factors** | **CVD Mortality / # of Participants, (%)** | | **Minimally Adjusted ^\|\|^** | | | **Fully Adjusted ^#^** | | |
| --- | --- | --- | --- | --- | --- | --- | --- | --- |
|  | **Men** | **Women** | **Men** | **Women** | **P-value** | **Men** | **Women** | **P-value** |
| **Social determinants of health ^†^** | | | | | | | | |
| Employment status |  |  |  |  |  |  |  |  |
| Employed, student, or retired | 1,042/7,515 (13.9) | 795/6,922 (11.5) | Ref | Ref | <0.001 | Ref | Ref | 0.005 |
| Unemployed | 174/1,264 (13.8) | 205/2,071 (9.9) | 2.36 (1.94, 2.89) | 1.32 (1.09, 1.60) |  | 1.70 (1.36, 2.12) | 1.11 (0.91, 1.36) |  |
| Family income-to-poverty ratio |  |  |  |  |  |  |  |  |
| ≥300% | 324/2,900 (11.2) | 193/2,497 (7.7) | Ref | Ref | 0.93 | Ref | Ref | 0.85 |
| <300% | 804/5,029 (16.0) | 682/5,476 (12.5) | 1.61 (1.37, 1.90) | 1.63 (1.36, 1.96) |  | 1.27 (1.05, 1.53) | 1.24 (1.02, 1.50) |  |
| Food security |  |  |  |  |  |  |  |  |
| Full security | 1,001/6,950 (14.4) | 815/6,907 (11.8) | Ref | Ref | 0.71 | Ref | Ref | 0.98 |
| Marginal, low, or very low security | 191/1,646 (11.6) | 158/1,886 (8.4) | 1.60 (1.29, 1.98) | 1.51 (1.23, 1.86) |  | 1.07 (0.88, 1.31) | 1.08 (0.85, 1.37) |  |
| Education level |  |  |  |  |  |  |  |  |
| High school graduate or higher | 705/5,715 (12.3) | 550/5,854 (9.4) | Ref | Ref | 0.72 | Ref | Ref | 0.74 |
| Less than high school | 506/3,046 (16.6) | 443/3,115 (14.2) | 1.44 (1.21, 1.71) | 1.39 (1.19, 1.61) |  | 1.08 (0.90, 1.31) | 1.04 (0.88, 1.23) |  |
| Regular health care access |  |  |  |  |  |  |  |  |
| At least one regular health care facility | 1,088/7,623 (14.3) | 929/8,251 (11.3) | Ref | Ref | 0.29 | Ref | Ref | 0.23 |
| None or emergency room | 128/1,160 (11.0) | 70/749 (9.3) | 1.07 (0.86, 1.34) | 1.30 (0.98, 1.72) |  | 0.98 (0.79, 1.21) | 1.24 (0.90, 1.71) |  |
| Health insurance status |  |  |  |  |  |  |  |  |
| Private | 592/4,418 (13.4) | 462/4,433 (10.4) | Ref | Ref | 0.72 | Ref | Ref | 1.00 |
| Government or none | 608/4,302 (14.1) | 521/4,496 (11.6) | 1.27 (1.11, 1.45) | 1.23 (1.05, 1.44) |  | 1.04 (0.90, 1.21) | 1.04 (0.87, 1.24) |  |
| Home ownership |  |  |  |  |  |  |  |  |
| Own home | 912/6,699 (13.6) | 695/6,560 (10.6) | Ref | Ref | 0.11 | Ref | Ref | 0.15 |
| Rent home or other arrangement | 291/1,955 (14.9) | 287/2,295 (12.5) | 1.60 (1.32, 1.92) | 1.31 (1.12, 1.53) |  | 1.22 (0.99, 1.52) | 1.00 (0.84, 1.19) |  |
| Number of persons per room in home |  |  |  |  |  |  |  |  |
| ≤1 person | 1,181/8,312 (14.2) | 948/8,504 (11.1) | Ref | Ref | 0.25 | Ref | Ref | 0.26 |
| >1 person | 21/318 (6.6) | 29/334 (8.7) | 0.74 (0.40, 1.38) | 1.16 (0.72, 1.85) |  | 0.59 (0.32, 1.10) | 0.95 (0.57, 1.59) |  |
| Marital status |  |  |  |  |  |  |  |  |
| Married or living with a partner | 788/6,205 (12.7) | 300/3,934 (7.6) | Ref | Ref | 0.74 | Ref | Ref | 0.69 |
| Not married nor living with a partner | 416/2,523 (16.5) | 676/4,959 (13.6) | 1.52 (1.33, 1.74) | 1.47 (1.26, 1.71) |  | 1.21 (1.05, 1.40) | 1.16 (0.98, 1.36) |  |
| **Behavioral factors ^‡^** | | | | | | | | |
| Smoking |  |  |  |  |  |  |  |  |
| Former or never smoking | 1,031/7,396 (13.9) | 907/8,098 (11.2) | Ref | Ref | 0.44 | Ref | Ref | 0.78 |
| Current smoking | 183/1,375 (13.3) | 90/889 (10.1) | 2.01 (1.63, 2.49) | 1.75 (1.31, 2.34) |  | 1.70 (1.37, 2.12) | 1.61 (1.17, 2.23) |  |
| Alcohol drinking |  |  |  |  |  |  |  |  |
| Moderate or no alcohol drinking | 1,048/7,496 (14.0) | 863/7,854 (11.0) | Ref | Ref | 0.09 | Ref | Ref | 0.46 |
| Heavy alcohol drinking | 85/695 (12.2) | 40/429 (9.3) | 0.93 (0.70, 1.24) | 0.61 (0.42, 0.89) |  | 0.94 (0.71, 1.26) | 0.79 (0.53, 1.16) |  |
| Diet quality |  |  |  |  |  |  |  |  |
| HEI ≥52 | 577/4,297 (13.4) | 521/5,027 (10.4) | Ref | Ref | 0.56 | Ref | Ref | 0.39 |
| HEI <52 | 462/3,157 (14.6) | 321/2,678 (12.0) | 1.26 (1.06, 1.51) | 1.18 (1.01, 1.38) |  | 1.07 (0.89, 1.28) | 0.96 (0.80, 1.15) |  |
| Leisure-time physical activity |  |  |  |  |  |  |  |  |
| Any | 451/3,774 (12.0) | 247/3,266 (7.6) | Ref | Ref | 0.19 | Ref | Ref | 0.08 |
| No | 764/4,999 (15.3) | 752/5,727 (13.1) | 1.65 (1.43, 1.90) | 1.93 (1.63, 2.30) |  | 1.23 (1.05, 1.45) | 1.56 (1.31, 1.85) |  |
| Sleep duration |  |  |  |  |  |  |  |  |
| 6-8 hours per day | 430/4,383 (9.8) | 296/4,326 (6.8) | Ref | Ref | 0.12 | Ref | Ref | 0.16 |
| <6 or >8 hours per day | 182/1,921 (9.5) | 180/2,125 (8.5) | 1.08 (0.88, 1.32) | 1.33 (1.11, 1.58) |  | 1.01 (0.84, 1.22) | 1.23 (1.01, 1.49) |  |
| Depression |  |  |  |  |  |  |  |  |
| PHQ-9 <10 | 511/5,397 (9.5) | 345/5,255 (6.6) | Ref | Ref | 0.85 | Ref | Ref | 0.90 |
| PHQ-9 ≥10 | 35/352 (9.9) | 43/551 (7.8) | 1.47 (0.99, 2.18) | 1.40 (1.03, 1.90) |  | 1.09 (0.76, 1.57) | 1.12 (0.80, 1.57) |  |
| **Metabolic factors ^§^** | | | | | | | | |
| Obesity |  |  |  |  |  |  |  |  |
| No | 791/5,742 (13.8) | 572/5,217 (11.0) | Ref | Ref | 0.33 | Ref | Ref | 0.55 |
| Yes | 336/2,729 (12.3) | 343/3,487 (9.8) | 1.40 (1.19, 1.66) | 1.26 (1.09, 1.45) |  | 1.19 (0.96, 1.47) | 1.09 (0.91, 1.30) |  |
| Central obesity |  |  |  |  |  |  |  |  |
| No | 485/3,717 (13.0) | 202/1,846 (10.9) | Ref | Ref | 0.34 | Ref | Ref | 0.54 |
| Yes | 599/4,402 (13.6) | 630/6,311 (10.0) | 1.24 (1.08, 1.41) | 1.12 (0.96, 1.30) |  | 1.00 (0.84, 1.19) | 0.91 (0.73, 1.13) |  |
| Hypertension |  |  |  |  |  |  |  |  |
| No | 203/1,917 (10.6) | 109/1,602 (6.8) | Ref | Ref | 0.94 | Ref | Ref | 0.48 |
| Yes | 978/6,673 (14.7) | 865/7,199 (12.0) | 1.34 (1.09, 1.65) | 1.36 (1.09, 1.68) |  | 1.17 (0.95, 1.44) | 1.05 (0.83, 1.33) |  |
| Diabetes |  |  |  |  |  |  |  |  |
| No | 807/6,089 (13.3) | 694/6,543 (10.6) | Ref | Ref | 0.77 | Ref | Ref | 0.98 |
| Yes | 409/2,695 (15.2) | 306/2,457 (12.5) | 1.84 (1.57, 2.16) | 1.78 (1.53, 2.06) |  | 1.42 (1.20, 1.68) | 1.42 (1.18, 1.70) |  |
| High cholesterol |  |  |  |  |  |  |  |  |
| No | 881/6,502 (13.5) | 752/7,154 (10.5) | Ref | Ref | 0.10 | Ref | Ref | 0.19 |
| Yes | 254/1,780 (14.3) | 164/1,201 (13.7) | 1.11 (0.92, 1.33) | 1.37 (1.14, 1.64) |  | 1.00 (0.83, 1.21) | 1.19 (0.98, 1.46) |  |
| Albumin-to-creatinine ratio |  |  |  |  |  |  |  |  |
| <30 mg/g | 742/6,469 (11.5) | 585/6,853 (8.5) | Ref | Ref | 0.16 | Ref | Ref | 0.17 |
| ≥30 mg/g | 407/2,014 (20.2) | 326/1,737 (18.8) | 2.18 (1.84, 2.57) | 2.54 (2.17, 2.97) |  | 1.71 (1.43, 2.04) | 2.02 (1.71, 2.40) |  |
| Estimated glomerular filtration rate |  |  |  |  |  |  |  |  |
| ≥60 ml/min/1.73 m^2^ | 763/6,572 (11.6) | 533/6,416 (8.3) | Ref | Ref | 0.77 | Ref | Ref | 0.77 |
| <60 ml/min/1.73 m^2^ | 371/1,687 (22.0) | 381/1,914 (19.9) | 1.68 (1.44, 1.96) | 1.73 (1.48, 2.04) |  | 1.40 (1.20, 1.63) | 1.45 (1.21, 1.74) |  |

Abbreviations: CI, confidence interval; HEI, healthy eating index; PHQ-9, Patient Health Questionnaire-9; NHANES, National Health and Nutrition Examination Survey.

^*^ CVD mortality/number of participants is unweighted using original data. All the other numbers in the table are weighted accounting for the complex, multistage, probability design of NHANES and multiple imputation. For Cox regression, age is treated as timescale and models are stratified by birth cohort.

^†^ Employment is defined as at work, student or retired. Food insecurity is assessed with the 18-item US Food Security Survey Module with zero affirmative responses indicating high food security and ≥1 affirmative responses indicating marginal or lower food security.

^‡^ Heavy alcohol drinking is defined with more than 14 (7) drinks per week for men (women); The Healthy Eating Index (HEI) uses a scoring system to assess how well a set of foods aligns with key recommendations of the Dietary Guidelines for Americans. The scores range from 0 to 100 with a higher score indicating greater consistency of the diet with the Dietary Guidelines for Americans. Median HEI score was 52 among NHANES 1999-2018 participants; Depression was assessed with the Patient Health Questionnaire-9 (PHQ-9) with scores ≥10 indicating depression.

^§^ Obesity is defined as body mass index ≥30 kg/m2; central obesity is defined as waist circumference ≥102 cm for men and ≥ 88 cm for women; hypertension is defined as systolic blood pressure ≥130 mmHg and/or diastolic blood pressure ≥80 mmHg or use of antihypertensive medications; and diabetes is defined as fasting glucose ≥126mg/dL or hemoglobin A1c ≥6.5% or diagnosed diabetes.

**^||^** Adjusted for sex and race. For Cox regression, age is treated as timescale and models are stratified by birth cohort.

**^#^** Adjusted for sex, race, and all other psychosocial, behavioral, and metabolic risk factors listed in the table. For Cox regression, age is treated as timescale and models are stratified by birth cohort.

**Supplemental Table 6. Population Attributable Fractions for Cardiovascular Diseases Mortality Among US Adults by Sex and Age Group ^*^**

| **Risk Factors** | **Population Attributable Fraction, % (95% CI)** | | | | | |
| --- | --- | --- | --- | --- | --- | --- |
|  | **All US Adults ^†^** | | **Age <60 years ^‡^** | | **Age ≥60 years ^§^** | |
|  | **Men** | **Women** | **Men** | **Women** | **Men** | **Women** |
| **Social Determinants of Health** | | | | | | |
| Unemployed | 4.9 (2.5 - 7.2) | -- | 13.1 (4.4 - 21.9) | -- | 3.2 (1.4 - 4.9) | -- |
| Family income-to-poverty ratio <300% | 8.7 (2.8 - 14.6) | 10.0 (2.8 - 17.2) | -- | -- | 9.6 (3.0 - 16.1) | 8.2 (0.3 - 16.1) |
| Marginal or lower food security | -- | -- | -- | -- | -- | -- |
| Less than high school education | -- | -- | -- | -- | -- | -- |
| No regular health care access | -- | -- | -- | -- | -- | -- |
| No private health insurance | -- | -- | -- | -- | 1.4 (-2.5, 5.4) | -- |
| No home ownership | 2.5 (0.3 - 4.8) | -- | -- | -- | 2.3 (-0.1 - 4.6) | -- |
| >1 person per room in home | -- | -- | -- | -- | -- | -- |
| Not married nor living with a partner | 5.1 (2.1 - 8.1) | 6.1 (0.6 - 11.7) | 11.4 (1.7 - 21.1) | -- | 4.0 (1.4 - 6.5) | -- |
| **Behavioral Factors** | | | | | | |
| Current smoking | 4.1 (1.9 - 6.3) | 3.7 (1.8 - 5.6) | 8.8 (0.5 - 17.1) | 14.6 (4.7 - 24.6) | 3.4 (1.4 - 5.3) | 2.2 (0.3 - 4.1) |
| Heavy alcohol drinking | -- | -- | -- | -- | -- | -- |
| Unhealthy diet | -- | -- | -- | -- | -- | -- |
| Leisure-time physical inactivity | 6.6 (1.5 - 11.7) | 14.7 (7.7 - 21.7) | -- | -- | 8.1 (2.3 - 13.9) | 18.5 (11.5 - 25.6) |
| Sleep duration <6 or >8 hours/day | -- | 3.5 (-0.0 - 7.1) | -- | -- | -- | 4.7 (0.8 - 8.5) |
| Depression | -- | -- | -- | -- | -- | -- |
| **Metabolic Factors** | | | | | | |
| Obesity | -- | -- | -- | -- | -- | -- |
| Central obesity | -- | -- | -- | -- | -- | -- |
| Hypertension | 14.3 (6.8 - 21.8) | -- | 22.8 (9.0 - 36.7) | 24.3 (6.7 - 41.9) | -- | -- |
| Diabetes | 6.3 (2.7 - 9.9) | 6.5 (3.2 - 9.9) | 6.5 (-1.2 - 14.2) | 17.2 (6.0 - 28.3) | 6.9 (3.1 - 10.6) | 5.6 (2.2 - 9.1) |
| High cholesterol | -- | 1.7 (-0.0 - 3.4) | -- | -- | -- | -- |
| Albuminuria | 10.0 (6.5 - 13.5) | 11.7 (7.8 - 15.5) | 11.3 (4.6 - 17.9) | 8.9 (0.5 - 17.2) | 9.9 (6.4 - 13.4) | 12.2 (8.2 - 16.2) |
| eGFR <60 ml/min/1.73 m^2^ | 5.5 (2.6 - 8.3) | 8.9 (5.0 - 12.8) | 2.1 (-0.7 - 4.9) | 8.9 (1.8 - 15.9) | 6.9 (3.7 - 10.1) | 8.5 (3.7 - 13.2) |
| **Total** | 67.9 (62.4 - 73.4) | 66.8 (60.6 - 73.1) | 76.1 (67.8 - 84.3) | 73.9 (60.1 - 87.7) | 54.2 (47.7 - 60.6) | 59.9 (52.4 - 67.5) |

Abbreviations: CI, confidence interval; eGFR, estimated glomerular filtration rate.

^*^ Estimates were calculated accounting for the complex, multistage, probability design of the National Health and Nutrition Examination Survey and multiple imputation.

^†^ All risk factors were included in one model and additionally adjusted for survey year, age, race/ethnicity, food security, education level, regular healthcare access, health insurance, home crowdedness, alcohol drinking, diet quality, sleep duration, depression, obesity, central obesity, and high cholesterol for men; and survey year, age, race/ethnicity, employment, food security, home ownership, education level, regular healthcare access, health insurance, home crowdedness, alcohol drinking, diet quality, depression, obesity, central obesity and hypertension for women.

^‡^ All risk factors were included in one model and additionally adjusted for survey year, age, race/ethnicity, family income-to-poverty ratio, food security, home ownership, education level, regular healthcare access, health insurance, home crowdedness, alcohol drinking, diet quality, leisure-time physical activity, sleep duration, depression, obesity, central obesity, and high cholesterol for men; and survey year, age, race/ethnicity, for women.

^§^ All risk factors were included in one model and additionally adjusted for survey year, age, race/ethnicity, food security, education level, regular healthcare access, health insurance, home crowdedness, alcohol drinking, diet quality, sleep duration, depression, obesity, central obesity, hypertension, and high cholesterol for men; and survey year, age, race/ethnicity, employment, food security, home ownership, education level, regular healthcare access, health insurance, home crowdedness, marital status, alcohol drinking, diet quality, depression, obesity, central obesity, hypertension and high cholesterol for women.
